# Multi-scale analysis of the HIF1A hypoxia–inflammation axis in severe COVID-19

**DOI:** 10.64898/2026.10.23.26366304

**Authors:** Marko N. Živanović, Ivana Raković, Ana Mirić, Miloš Z. Milosavljević, Nevena Milivojević Dimitrijević, Nenad Filipović, Jelena Košarić, Katarina Virijević, Marija Branković, Tamara Mladenović, Biljana Popovska Jovičić

## Abstract

**Objective and design:** To test whether the hypoxia-inducible factor 1-alpha (HIF1A) axis links transcriptional, cellular, biomarker and host-genetic features of severe COVID-19, in a retrospective multi-scale observational study of public cohorts and one hospital cohort.

**Material or subjects:** 35,268 hospitalized patients (Mexican national registry); 1,100 samples from five bulk RNA-seq cohorts; 25,773 peripheral neutrophils from six individuals; 326 hospitalized patients (São Paulo, Brazil); 93 hospitalized patients (University Clinical Center Kragujevac, Serbia).

**Treatment:** None.

**Methods:** Within-cohort effect sizes for a 10-gene HIF1A panel; single-cell pathway scoring; logistic regression and gradient boosting with SHAP; TaqMan genotyping of HIF1A rs11549465 and rs41508050; cross-validation with bootstrap confidence intervals.

**Results:** HIF1A (mean Cohen’s d +0.39) and LDHA (+0.76) were higher in severe disease in all five cohorts. Neutrophils of all three patients who failed extubation scored higher than those of both weaned patients and over-expressed S100A8/A9. C-reactive protein dominated ICU prediction; laboratory variables raised the AUC from 0.636 to 0.794. In Kragujevac, a clinical model predicted severe outcome (AUC 0.84, 95% CI 0.75–0.92). rs11549465 CT/TT carriers had lower mortality (1/11 vs 28/78; p = 0.095).

**Conclusions:** HIF1A pathway activation is reproducible across cohorts and localizes to an alarmin-rich neutrophil state; host-genetic effects require larger cohorts.

## Introduction

The clinical course of SARS-CoV-2 infection remains heterogeneous. Patients with similar age and comorbidity profiles progress either to recovery or to acute respiratory distress syndrome (ARDS), multi-organ failure and death [1]. Risk tools built on demographics and comorbidities reach only modest discrimination, and routine laboratory markers such as C-reactive protein (CRP), D-dimer and lactate dehydrogenase (LDH) improve prediction without explaining the underlying biology [2, 3].

Hypoxia-inducible factor 1-alpha (HIF1A) is a candidate integrator of these signals. Stabilized by tissue hypoxia or by inflammatory pseudo-hypoxia, HIF1A drives glycolysis (LDHA, SLC2A1, PDK1), vascular responses (VEGFA) and iron handling, and it interacts with NF-κB signalling [4–7]. In myeloid cells HIF1A is required for inflammatory function [8], and in neutrophils it prolongs survival under hypoxia through NF-κB activity [9]. In COVID-19, HIF1A-dependent glycolysis sustains monocyte responses and viral replication [10], and HIF1A promotes SARS-CoV-2 infection and the inflammatory response [11].

Most studies of this axis examined one level of evidence: transcripts in small cohorts, biomarkers without molecular context, or single polymorphisms without functional context. Two missense variants in the oxygen-dependent degradation (ODD) domain of HIF1A, rs11549465 (Pro582Ser) and rs41508050 (Ala588Thr), alter HIF1A transactivation in reporter assays [12], yet they are absent from commercial COVID-19 host-genetic panels, which centre on viral-entry and interferon loci [13]. Our group previously characterized interferon-lambda and ACE2/TMPRSS2 variants in an independent COVID-19 cohort at the same clinical center [14–16].

Here we combine four independent scales: population (Scale A), transcriptomic including single-cell data (Scale B), clinical biomarker (Scale C) and clinical-genetic (Scale D). We tested four hypotheses: (H1) demographic and comorbidity variables set a baseline level of discrimination; (H2) inflammatory and organ-dysfunction biomarkers add discrimination beyond that baseline; (H3) the HIF1A ODD-domain variants are associated with outcome (exploratory); and (H4) HIF1A pathway transcripts are consistently up-regulated in severe disease and localize to a cellular state associated with clinical course.

## Materials and methods

### Study design, ethics and reporting

This is a retrospective multi-scale observational study. Scale D used prospectively collected clinical data and blood samples from patients hospitalized at the University Clinical Center Kragujevac, Serbia, within the study “Investigation of the humoral response and of phenotypic and functional features of peripheral blood cells in patients with COVID-19”, approved by the Ethics Committee of the Clinical Center Kragujevac (No. 01/20-498, 5 May 2020). The study was conducted in accordance with the Declaration of Helsinki, and all participants gave written informed consent. Scales A–C used de-identified public data released under licences that permit secondary analysis. Reporting of the prediction models follows TRIPOD [17]; the completed checklist is provided as Online Resource 3.

### Scale A: population registry (Mexico)

We used the open COVID-19 data of the Mexican Directorate General of Epidemiology [18] (approximately 263,000 records). We retained patients with PCR-confirmed SARS-CoV-2 infection who were hospitalized (N ≈ 62,000) and had complete data for the primary variables (N = 35,268). Predictors were age, sex, radiographically confirmed pneumonia, diabetes, hypertension, chronic obstructive pulmonary disease (COPD), chronic kidney disease and smoking. Outcomes were intensive care unit (ICU) admission and death. Logistic regression (LR), random forest (RF) and XGBoost models were trained with 5-fold stratified cross-validation and evaluated on a held-out test set (20%; N = 7,054 for ICU, N = 7,057 for death), with 1,000 bootstrap replicates for confidence intervals. A model without the pneumonia variable quantified the contribution of radiographic findings. The filter cascade is given in Online Resource 1 (Section S1.1).

### Scale B: bulk transcriptomic meta-analysis

Five GEO cohorts were analysed: GSE152075 (N = 484), GSE157103 (N = 123), GSE171110 (N = 54), GSE212861 (N = 93) and GSE300696 (N = 346); total N = 1,100 [19–23]. We extracted ten HIF1A pathway genes: HIF1A, VEGFA, SLC2A1, LDHA, PDK1, IL6, TNF, CXCL8, STAT3 and MMP9. Raw expression differed up to 1,000-fold between cohorts, and class labels were perfectly aligned with cohort (a control group existed only in GSE152075). We therefore did not pool the cohorts. Within each cohort, genes were z-standardized, a multivariable LR model on the 10-gene panel was evaluated with 5-fold cross-validation, and effect sizes were expressed as Cohen’s d per gene and cohort.

### Scale B: single-cell transcriptomics of neutrophils

Dataset GSE234904 [24] contains 10x Chromium profiles of peripheral blood neutrophils from six individuals: one healthy donor, two patients with COVID-19 ARDS weaned from ventilation within 28 days (Weaned) and three who were not (NonWeaned). Cells with fewer than 200 detected genes or more than 20% mitochondrial reads were removed, giving 25,773 cells and 8,050 genes. Sequencing depth differed between samples: median unique molecular identifier (UMI) counts per cell were 366– 608 in the three NonWeaned samples and 740–1,152 in the two Weaned samples. Data were normalized (10,000 counts per cell, log1p), 2,000 highly variable genes were selected per sample, and principal components were integrated with Harmony using sample as the batch key [25], followed by Leiden clustering (resolution 0.5) in Scanpy [26]. Five panel genes were reliably detected in neutrophils (HIF1A, LDHA, STAT3, MMP9, PDK1) and were combined into a per-cell pathway score (score_genes). Because cells are nested within patients, the primary comparison of NonWeaned and Weaned groups was the patient-level mean score; cell-level Wilcoxon tests and differential expression (Benjamini–Hochberg adjusted) are reported descriptively.

### Scale C: clinical biomarker cohort (Brazil)

The Sírio-Libanês Hospital dataset [27] contains 1,925 serial observations of 385 hospitalized patients in five time windows after admission. To predict early ICU transfer, we used variables from the first 4 h as input and defined the outcome as ICU admission in any later window. Patients admitted directly to the ICU were excluded to avoid leakage, leaving 326 patients and 30 variables. The data were split 80/20 stratified by outcome (training N = 260, test N = 66). LR, RF (300 trees, maximum depth 10), XGBoost (300 trees, maximum depth 5, learning rate 0.05) and LightGBM were trained; XGBoost was additionally Platt-calibrated. Feature contributions were estimated with SHAP values. AUCs carry 1,000-replicate bootstrap confidence intervals, and clinical utility was assessed by decision-curve analysis.

### Scale D: clinical-genetic cohort (Kragujevac)

Patients with PCR-confirmed SARS-CoV-2 infection hospitalized at the University Clinical Center Kragujevac were enrolled from May 2020. Of 100 enrolled patients, 93 had complete clinical and genotype data. Recorded variables were age, sex, comorbidities (diabetes, hypertension, COPD/asthma, neurological disease, malignancy, chronic renal insufficiency), COVID-19 vaccination, chest X-ray score, oxygen saturation (SpO_2_), arterial pO_2_ and admission laboratory values (Table 2). Two outcomes were defined: in-hospital mortality, used to describe the cohort and to test genotype associations, and a composite severe outcome (high-flow oxygen therapy, mechanical ventilation or death; 30 events), used for the prediction model.

**Table 1.**
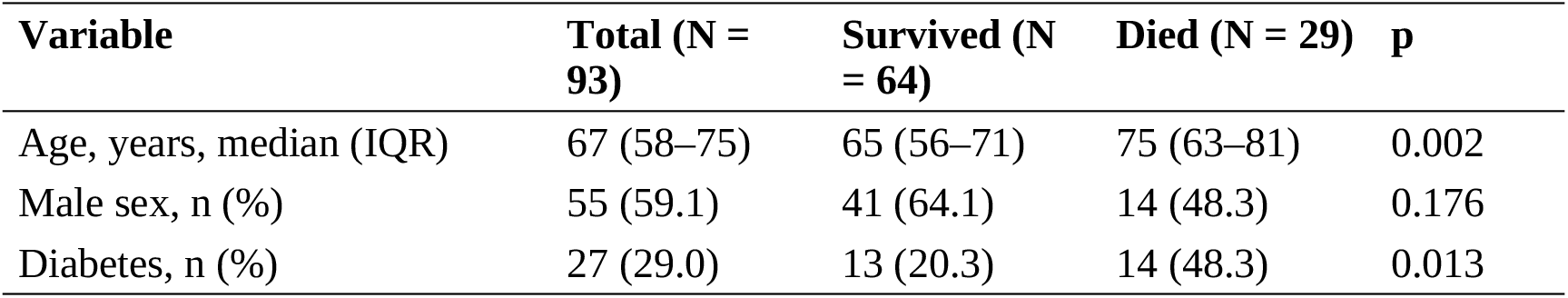
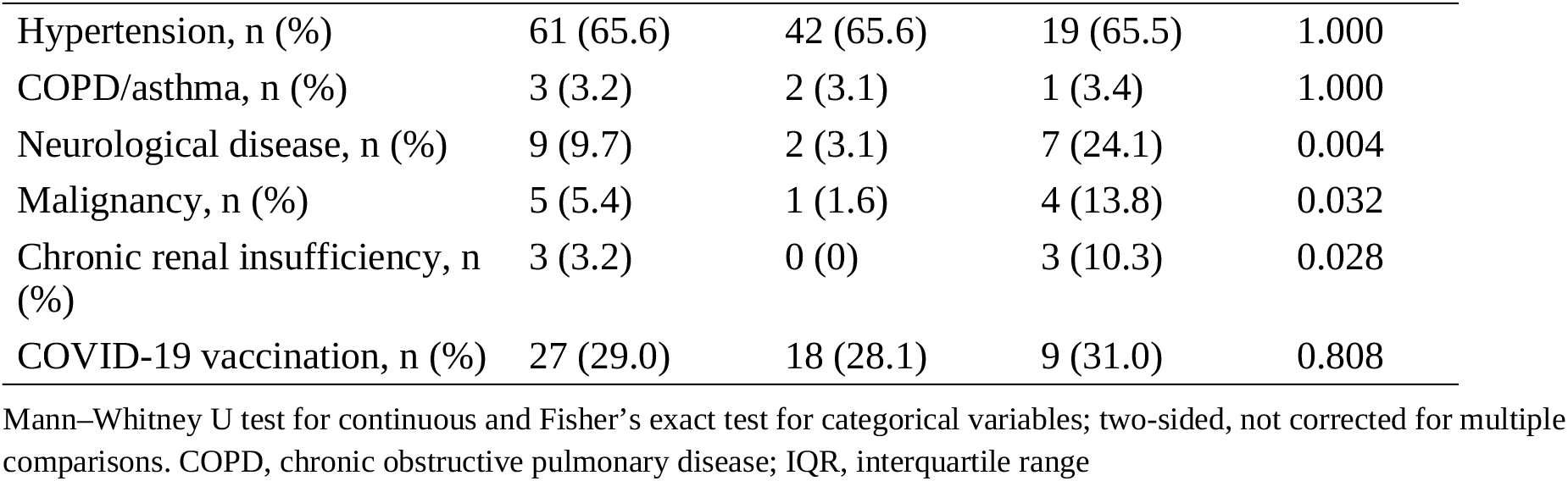
Demographic and clinical characteristics of the Kragujevac cohort by in-hospital outcome.

**Table 2.** Admission laboratory and radiological parameters by in-hospital outcome, median (IQR)

| Parameter | Total (N = 93) | Survived (N = 64) | Died (N = 29) | p |
| --- | --- | --- | --- | --- |
| Chest X-ray score | 8 (7–12) | 8 (6–9) | 14 (12–16) | <0.001 |
| SpO<sub>2</sub> (%) | 90 (86.2–92) | 92 (88.8–93) | 86 (77.2–90) | <0.001 |
| pO<sub>2</sub> (kPa) | 7.5 (6.3–8.0) | 7.5 (6.8–8.1) | 6.2 (5.4–7.7) | 0.003 |
| Leukocytes (×10<sup>9</sup>/L) | 8.5 (6.1–10.9) | 8.5 (5.9–10.3) | 9.2 (6.9–12.2) | 0.274 |
| Lymphocytes (×10<sup>9</sup>/L) | 0.7 (0.6–0.9) | 0.7 (0.6–0.9) | 0.6 (0.5–1.1) | 0.937 |
| Platelets (×10<sup>9</sup>/L) | 208 (160–268) | 222 (175–288) | 176 (134–247) | 0.011 |
| CRP (mg/L) | 90.5 (50.6–146.4) | 88.8 (41.6–129.5) | 94.3 (80.6–171.5) | 0.143 |
| LDH (U/L) | 757 (611–928) | 732 (593–899) | 999 (648–1200) | 0.005 |
| D-dimer (mg/L) | 1.0 (0.6–1.9) | 0.8 (0.6–1.6) | 1.2 (0.8–3.2) | 0.027 |
| Ferritin (µg/L) | 899 (446–1483) | 853 (420–1313) | 965 (461–1651) | 0.353 |
| IL-6 | 70 (32.6–102) | 61.6 (32.5–89.9) | 80 (39.4–174.2) | 0.087 |
| Urea (mmol/L) | 7.4 (5.5–10.8) | 6.9 (5.5–9.6) | 9.7 (6.7–16.3) | 0.006 |
| Creatinine (µmol/L) | 87 (73–114) | 82 (72–103) | 110 (80–156) | 0.006 |
| AST (U/L) | 41 (28.5–60) | 38 (28–49) | 53 (38.5–65.8) | 0.031 |
| Procalcitonin | 0.1 (0.1–0.2) | 0.1 (0–0.1) | 0.2 (0.1–0.3) | 0.012 |
| proBNP | 446.5 (157–1135) | 302 (117–835) | 610 (380–2237) | 0.023 |

Mann–Whitney U test; two-sided, not corrected for multiple comparisons. AST, aspartate aminotransferase; CRP, C-reactive protein; LDH, lactate dehydrogenase; proBNP, pro-B-type natriuretic peptide

The prediction model was an L2-penalized LR (C = 1, class-balanced weights) with 11 baseline predictors: age, sex, SpO_2_, pO_2_, diabetes, hypertension, COPD/asthma, neurological disease, malignancy, chronic renal insufficiency and vaccination. An aggregated comorbidity indicator was removed because of collinearity with hypertension; all remaining variance inflation factors were below 5. Missing values were imputed with the median and predictors were standardized; these parameters were estimated on the full cohort before cross-validation. Performance was estimated by leave-one-out cross-validation (LOOCV) with 1,000 bootstrap replicates for the confidence interval, and the operating threshold was chosen by the Youden index. With about 2.7 events per predictor, the model is exploratory. HIF1A genotypes were not entered into the prediction model; their association with mortality was tested separately. We also applied the Scale C model to the Kragujevac cohort as an external transfer test.

### DNA extraction and HIF1A genotyping

Genomic DNA was extracted from 200 μL of EDTA-anticoagulated whole blood with the PureLink™ Genomic DNA Mini Kit (Cat. No. K182002, Invitrogen, Thermo Fisher Scientific, Waltham, MA, USA). DNA concentration and purity were measured by UV spectrophotometry (NanoDrop, Thermo Fisher Scientific); samples with A260/A280 between 1.75 and 2.00 were genotyped. Two ODD-domain variants were genotyped by TaqMan allelic discrimination: rs11549465 (c.1744C>T, p.Pro582Ser; assay C 25473074_10) and rs41508050 (c.1762G>A, p.Ala588Thr; assay C 86499352_10), using TaqMan™ Genotyping Master Mix (Cat. No. 4371355, Applied Biosystems, Thermo Fisher Scientific) in 20 μL reactions on a Mic qPCR Cycler (Bio Molecular Systems, Upper Coomera, Australia) with BMS Workbench v2.0. Cycling was 95 °C for 10 min, then 40 cycles of 95 °C for 15 s and 60 °C for 60 s. Each plate included no-template controls, and all samples were run in technical duplicate; inter-run concordance was 97.6% (81/83 pairs). Reporting follows MIQE principles [28]. Assay details, Hardy–Weinberg equilibrium and reference-population comparison are given in Online Resource 1 (Section S1.5, Tables S1–S3). The same workflow was used in our earlier studies [14–16].

### Statistical analysis

Continuous variables are given as median (interquartile range, IQR) and categorical variables as n (%). Groups were compared with the Mann–Whitney U test or Fisher’s exact test. All tests were two-sided with α = 0.05. Comparisons in Tables 1 and 2 are descriptive and were not corrected for multiplicity. Genotype–mortality association was tested with Fisher’s exact test, with the conditional maximum-likelihood odds ratio (OR) and exact 95% confidence interval. Cohort size was fixed by enrolment, so no a priori power calculation was made; by simulation, the power to detect OR = 0.2 at the observed genotype frequency and baseline mortality was 34%. Analyses used Python 3.13 with scikit-learn, Scanpy 1.11.5, anndata 0.11.4, harmonypy, xgboost, lightgbm, statsmodels and scipy. Code, derived results and trained models are archived on Zenodo [29].

## Results

### Characteristics of the Kragujevac cohort

In-hospital mortality was 31.2% (29/93), and 30 patients (32.3%) reached the composite severe outcome. Patients who died were older (median 75 vs 65 years) and more often had diabetes, neurological disease, malignancy or chronic renal insufficiency (Table 1). They had lower SpO_2_, pO_2_ and platelet counts and higher chest X-ray scores, LDH, D-dimer, urea, creatinine, AST, procalcitonin and proBNP (Table 2). CRP, ferritin and IL-6 did not differ significantly. Treatments are listed in Online Resource 1 (Table S4).

### Scale A: demographic baseline

In 35,268 hospitalized patients, models restricted to demographic and comorbidity variables reached test-set AUCs of 0.661 (LR), 0.656 (RF) and 0.642 (XGBoost) for ICU admission, and 0.649 (LR) for death (Fig. 1). Radiographic pneumonia was the strongest predictor (OR ≈ 4.3), followed by age and diabetes (Online Resource 2, Figs. S8 and S9). Without the pneumonia variable the AUC fell to 0.609 (Online Resource 2, Fig. S10). Demographics and comorbidities therefore set a baseline near AUC 0.61–0.66 (H1).

**Fig 1.**
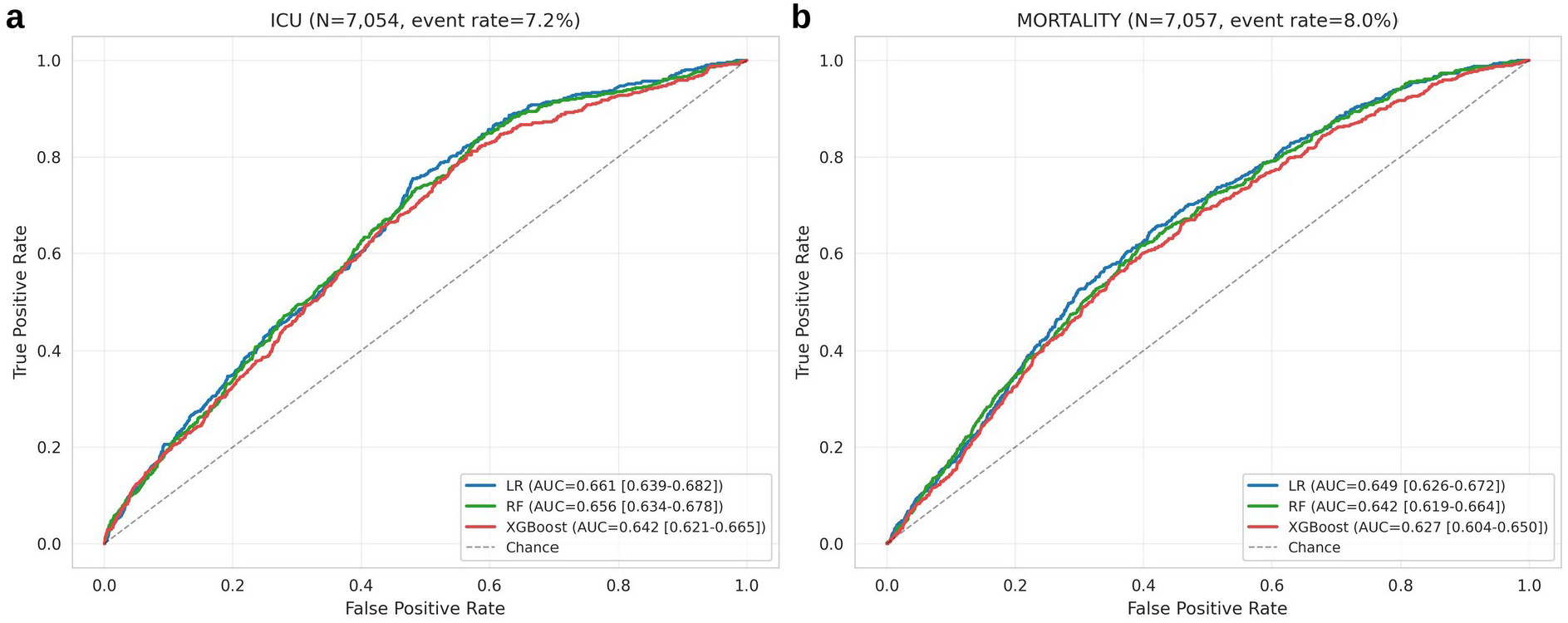
Scale A, receiver operating characteristic curves of logistic regression, random forest and XGBoost models based on demographic and comorbidity variables in hospitalized PCR-confirmed COVID-19 patients from the Mexican national registry (held-out test set). **a** ICU admission (N = 7,054). **b** Death (N = 7,057). AUC values with bootstrap 95% confidence intervals are given in the legends

### Scale B: convergent HIF1A pathway expression in bulk transcriptomes

Within-cohort LR models on the 10-gene panel reached AUCs of 0.72–0.99 across the five comparisons (Online Resource 1, Table S5; Online Resource 2, Fig. S11). HIF1A (mean d = +0.39) and LDHA (mean d = +0.76) were higher in the more severe group in all five cohorts, and STAT3 (+0.51), MMP9 (+0.49) and PDK1 (+0.22) in four of five (Fig. 2). TNF was lower in four cohorts (mean d = −0.19), and CXCL8 was heterogeneous, with d = −2.00 in GSE171110. Because panel genes moved in opposite directions, the unweighted mean of standardized genes discriminated poorly (AUC 0.45 vs 0.75 for LR in GSE152075).

**Fig 2.**
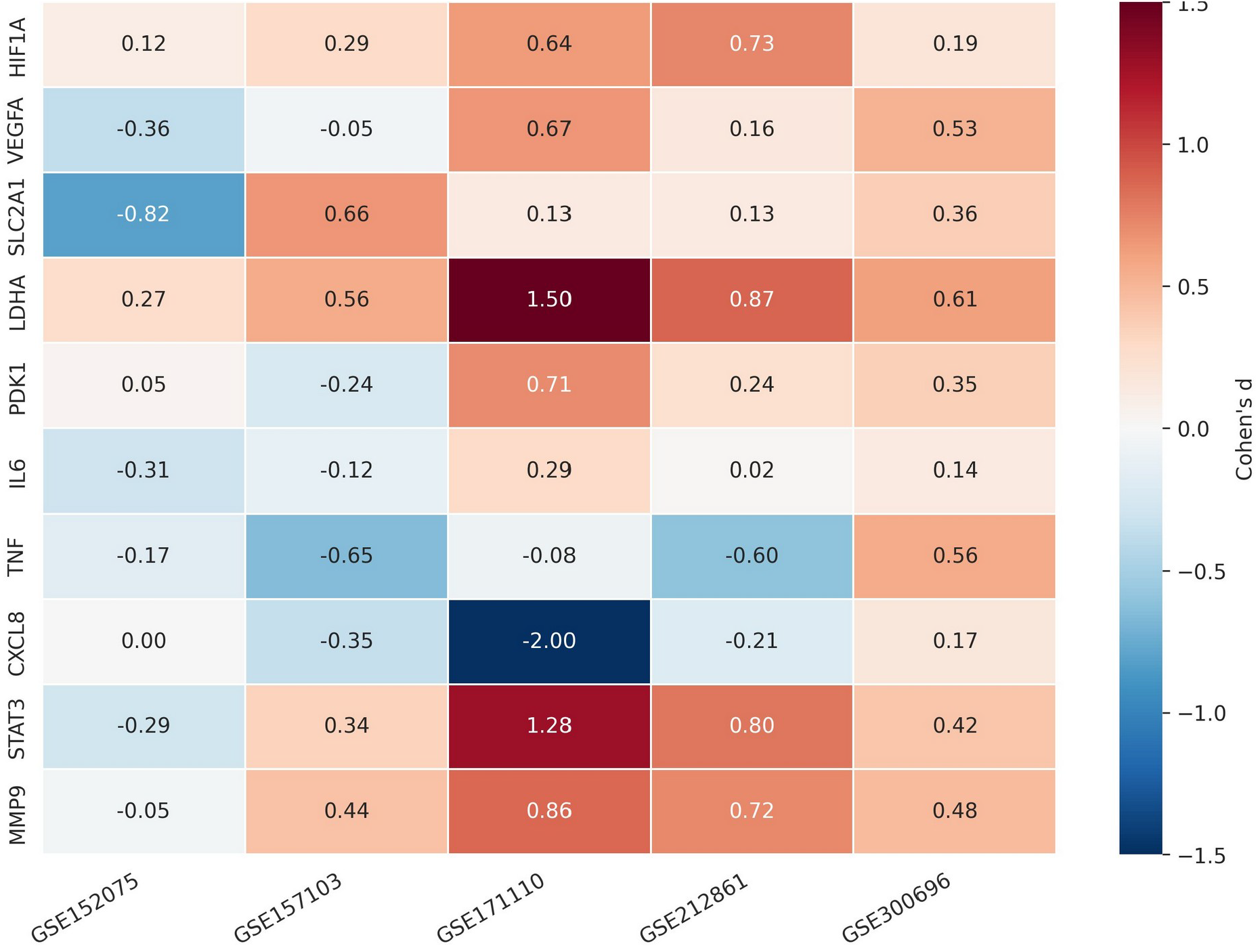
Scale B, Cohen’s d effect sizes of the ten HIF1A pathway genes in five GEO cohorts, each analysed within cohort. Red indicates higher expression in the more severe (positive) class and blue higher expression in the comparator class. Comparisons: GSE152075, COVID-19 vs control; GSE157103, ICU vs non-ICU; GSE171110, severe vs healthy; GSE212861, ARDS vs non-ARDS COVID-19; GSE300696, hospitalized vs convalescent

### Scale B: a HIF1A-high neutrophil state in patients who failed extubation

Harmony integration removed sample-dominated clusters and yielded eight Leiden clusters (Online Resource 2, Fig. S12). The HIF1A pathway score showed a gradient across the embedding (Fig. 3), with the highest mean score in cluster 2 (mean +0.25), and cluster 2 ranked highest within each patient group (Fig. 4e; Online Resource 2, Fig. S13). At patient level, the mean score of each of the three NonWeaned patients (+0.14, +0.08, +0.05) exceeded that of both Weaned patients (+0.02, −0.05) (Fig. 4c); at cell level the difference corresponded to d = +0.31. Scores did not differ between COVID-19 and the single healthy donor (d = +0.01), whose mean exceeded those of both Weaned patients.

**Fig 3.**
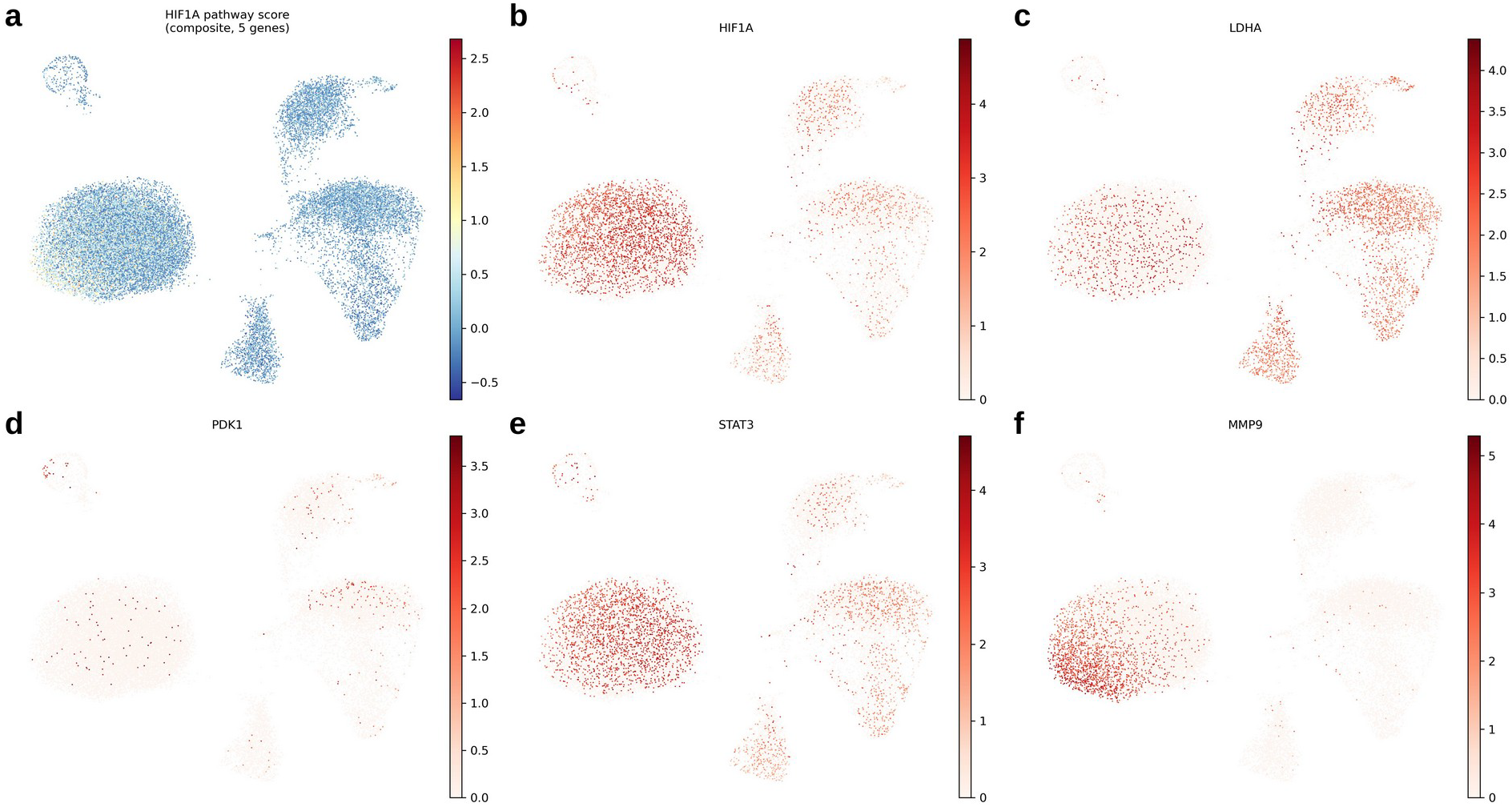
Scale B, single-cell RNA-seq of peripheral neutrophils (GSE234904; 25,773 cells, six individuals) on the Harmony-integrated UMAP embedding. **a** Composite HIF1A pathway score from the five genes detectable in neutrophils. **b–f** Log-normalized expression of HIF1A, LDHA, PDK1, STAT3 and MMP9

**Fig 4.**
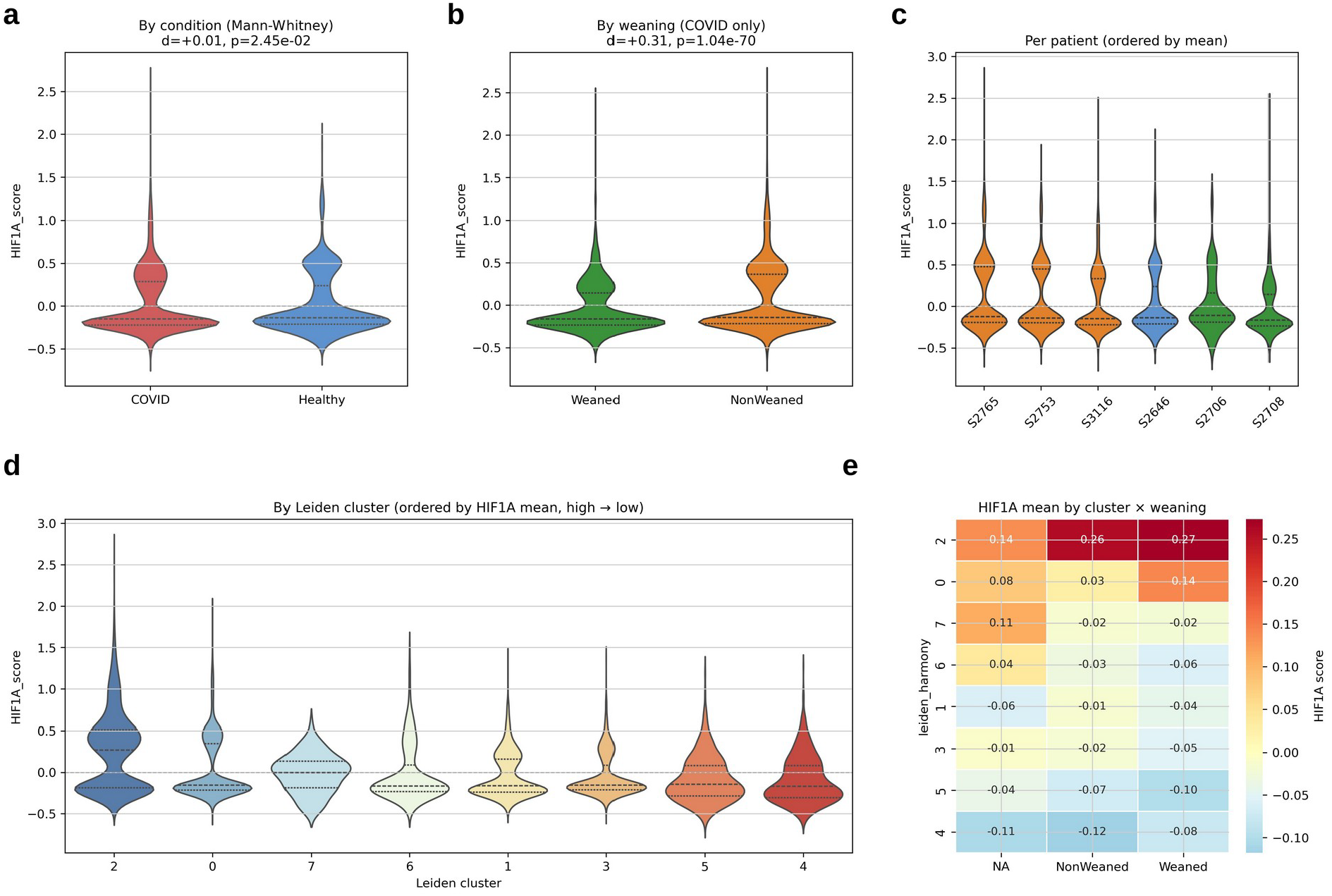
Scale B, distribution of the HIF1A pathway score. **a** COVID-19 vs healthy donor (one donor). **b** NonWeaned vs Weaned patients within COVID-19. **c** Per patient, ordered by mean score; orange, NonWeaned; green, Weaned; blue, healthy donor. **d** By Leiden cluster. **e** Mean score by cluster and weaning group (NA, healthy donor). Cell-level p values shown in the panel headers are descriptive because cells are nested within patients

Differential expression between NonWeaned and Weaned cells identified alarmins (S100A8, log_2_ fold change 8.5; S100A9, 6.1; S100A11, S100A6), interferon-induced transmembrane proteins (IFITM1, IFITM2, IFITM3), neutrophil activation genes (FCGR3B, MNDA, TYROBP) and iron and metal homeostasis genes (FTH1, MT2A) as higher in NonWeaned cells (Fig. 5). Fold changes of this size for transcripts that are already abundant in neutrophils should be read with the lower sequencing depth of the NonWeaned samples in mind (see Limitations).

**Fig 5.**
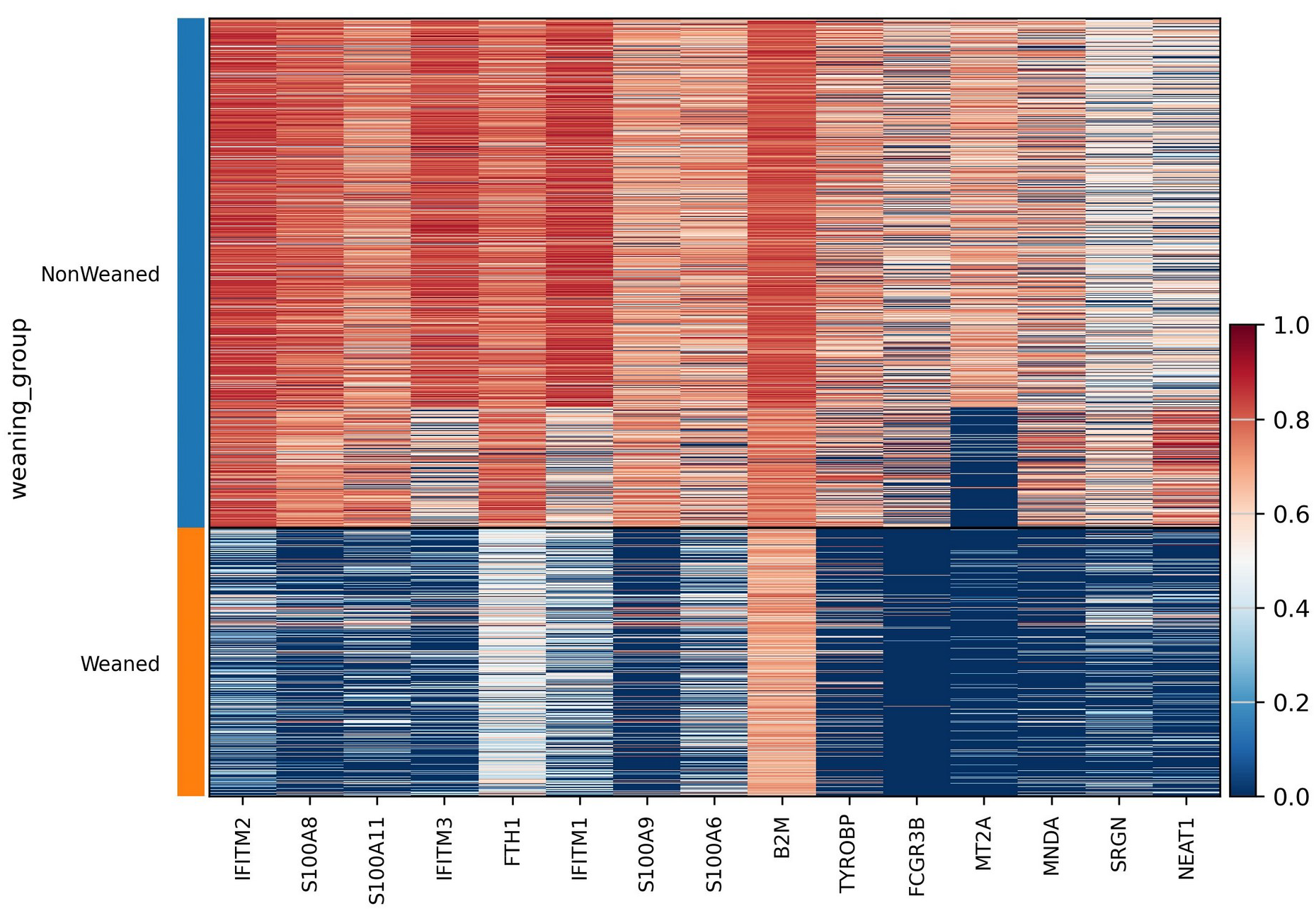
Scale B, top 15 differentially expressed genes between NonWeaned and Weaned COVID-19 neutrophils (Wilcoxon rank-sum test, Benjamini–Hochberg adjusted p < 0.001 for all genes; 22,766 cells). Rows are cells grouped by weaning status; colour shows scaled expression

### Scale C: inflammatory and organ-dysfunction markers add discrimination

In the Brazilian cohort, calibrated XGBoost predicted ICU transfer with a test-set AUC of 0.794 (95% CI 0.686–0.887) and a Brier score of 0.194, compared with 0.695 for LR (Online Resource 2, Fig. S14). CRP was the dominant feature (mean |SHAP| 0.77), followed by diastolic blood pressure (0.54), urea (0.50), age percentile (0.49), temperature (0.46) and creatinine (0.45) (Fig. 6; Online Resource 2, Fig. S15). A demographic baseline (age and sex) reached an AUC of 0.636 (95% CI 0.481–0.765), so admission laboratory variables added 0.158 (Fig. 7), supporting H2. Calibration reduced the Brier score from 0.209 to 0.194, and the calibrated model had a higher net benefit than treat-all and treat-none strategies at thresholds of 0.14–0.80 (Online Resource 2, Figs. S16 and S17).

**Fig 6.**
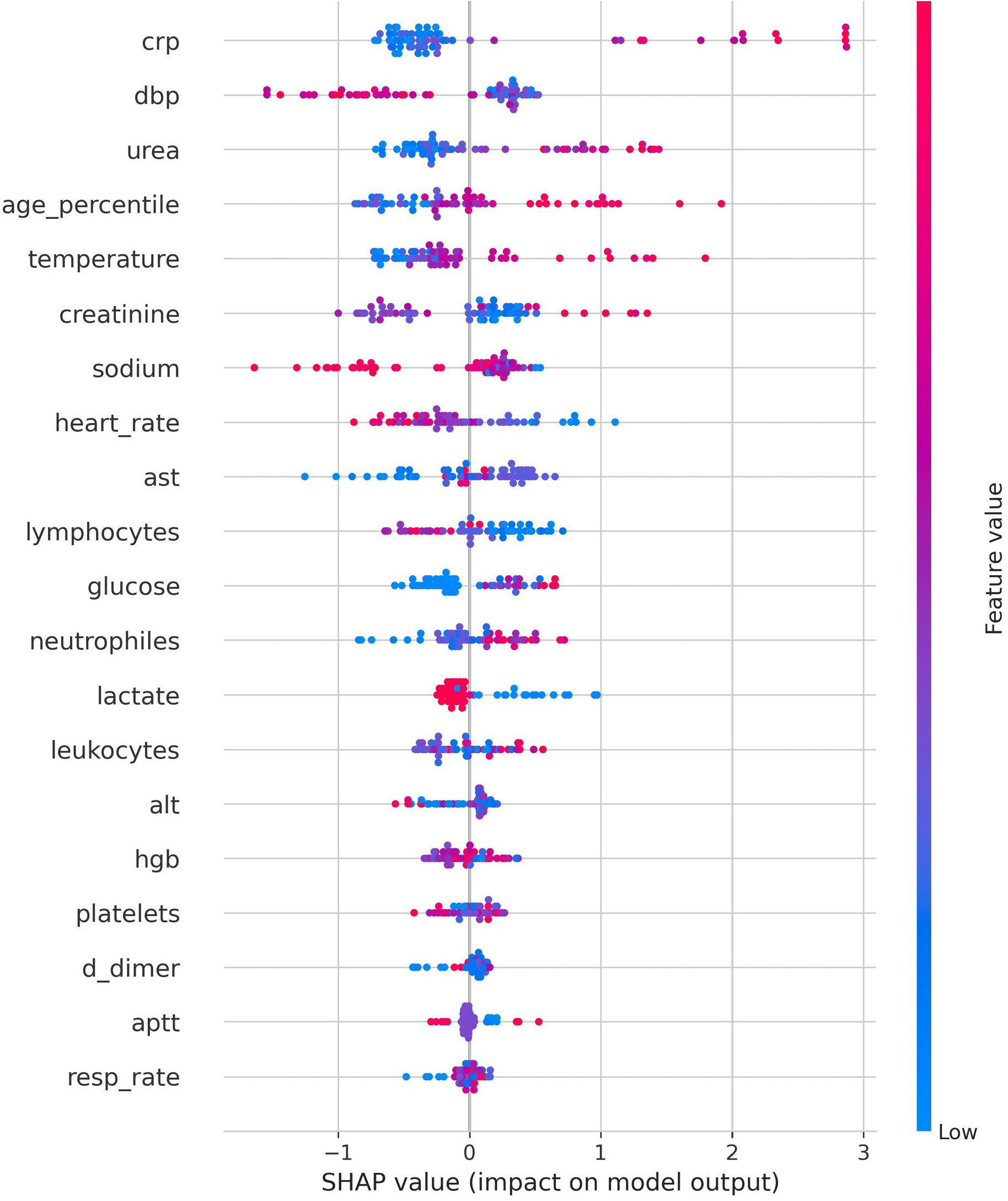
Scale C, SHAP summary of the calibrated XGBoost model for early ICU transfer in the Sírio-Libanês cohort. Each point is one patient; the horizontal position is the SHAP value (positive values push the prediction towards ICU transfer) and colour encodes the feature value (red, high; blue, low)

**Fig 7.**
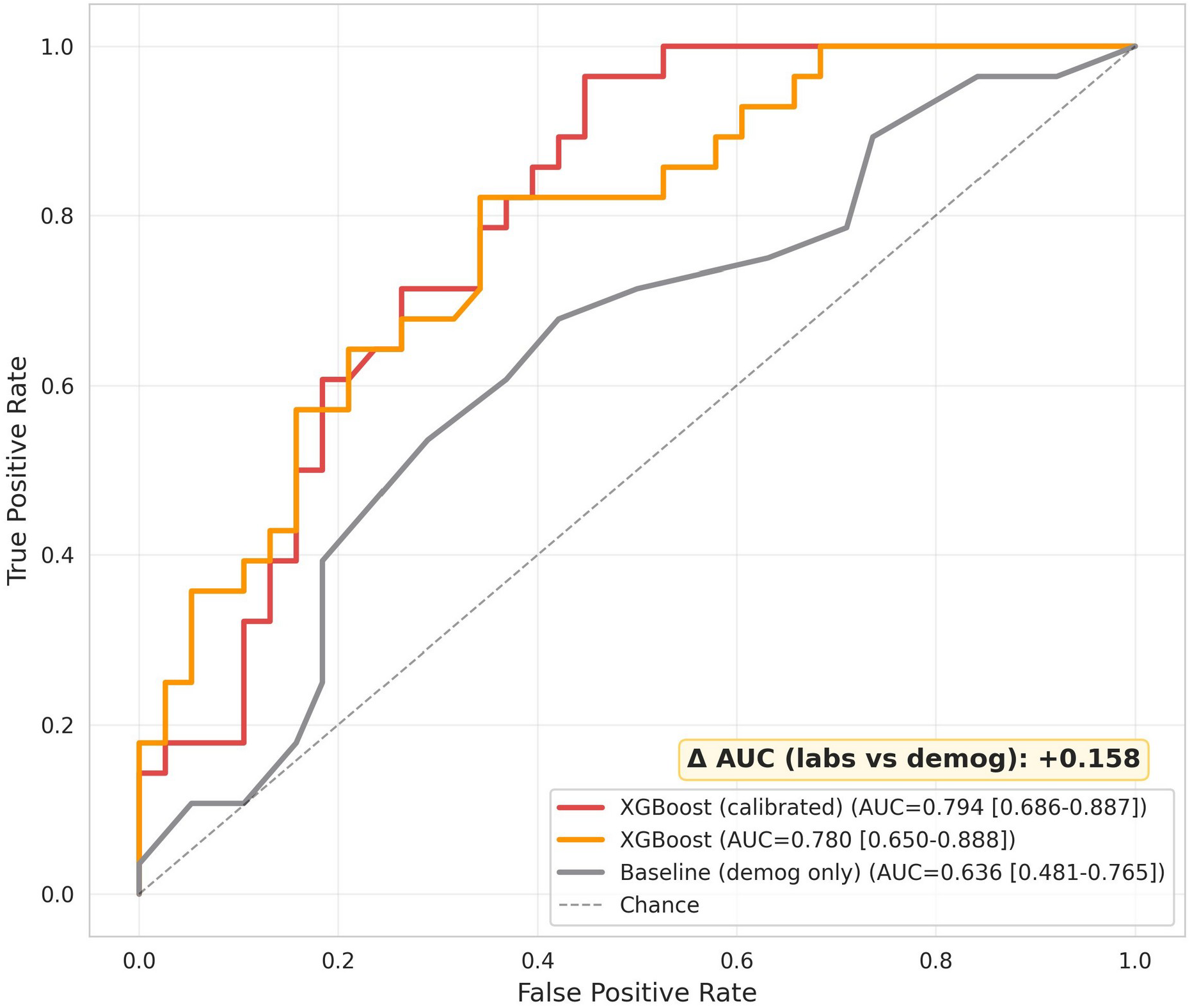
Scale C, incremental value of admission laboratory variables (test set, N = 66). Calibrated XGBoost with all variables (AUC 0.794), uncalibrated XGBoost (AUC 0.780) and a demographic baseline model (AUC 0.636). The difference between the full and baseline models is 0.158

### Scale D: clinical model and HIF1A genotypes in Kragujevac

The clinical model predicted the composite severe outcome with a LOOCV AUC of 0.840 (95% CI 0.751–0.916), sensitivity 0.87, specificity 0.73, negative predictive value 0.92 and Brier score 0.164 (Fig. 8). Diabetes, neurological disease, chronic renal insufficiency and age had the largest positive standardized coefficients, and hypertension had a negative coefficient (Table 3; Online Resource 2, Figs. S18–S20). A two-variable model with age and SpO_2_ alone reached an AUC of 0.797, so most discrimination came from age and oxygenation. Removing hypertension lowered the AUC by 0.026. The Scale C model transferred poorly to Kragujevac (AUC 0.50, 95% CI 0.36–0.63).

**Table 3.**
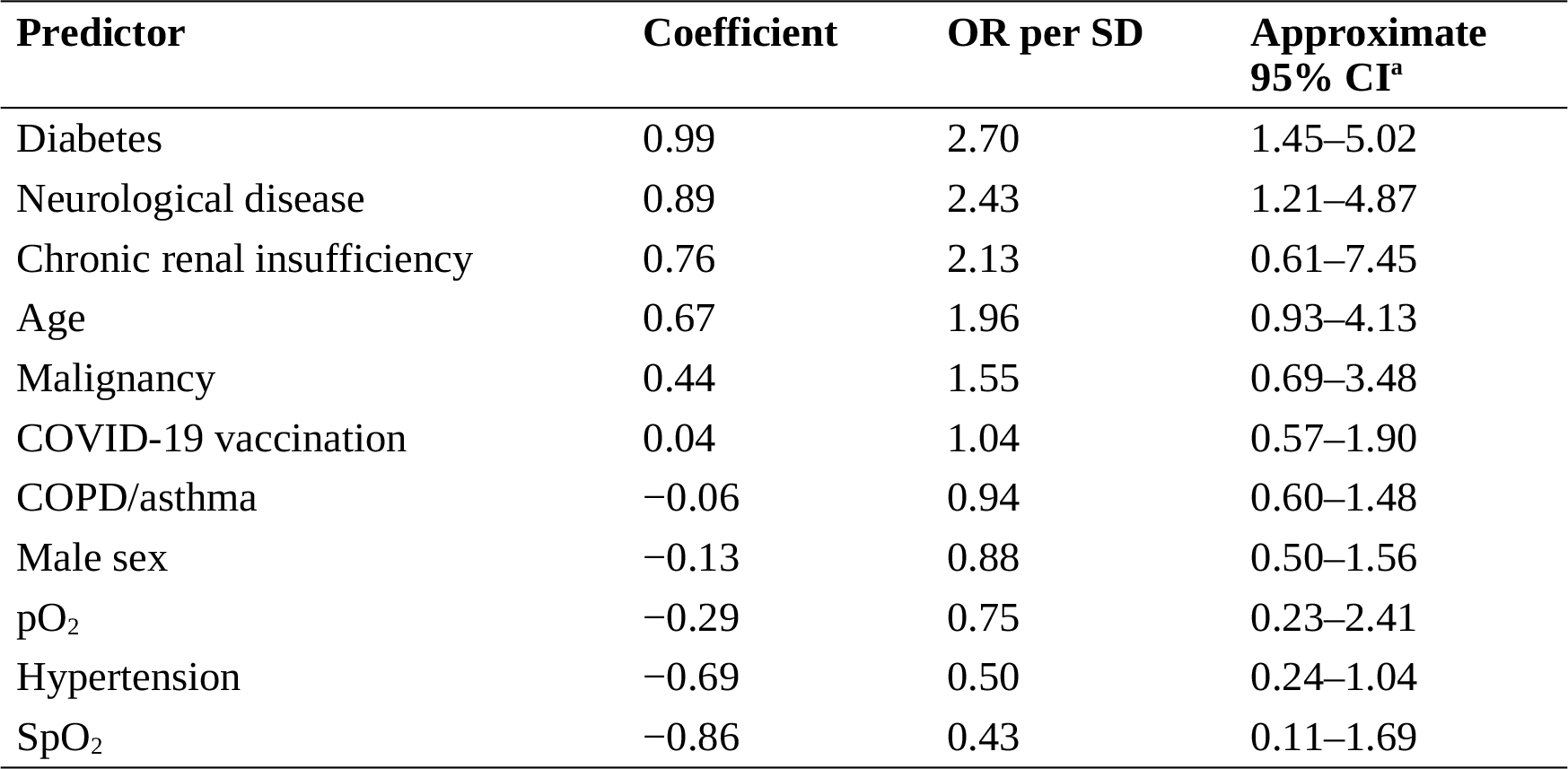
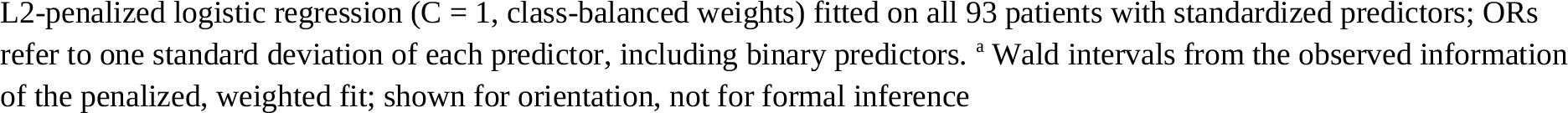
Standardized coefficients of the Kragujevac clinical model for the composite severe outcome.

| Predictor | Coefficient | OR per SD | Approximate 95% CI<sup>a</sup> |
| --- | --- | --- | --- |
| Diabetes | 0.99 | 2.70 | 1.45–5.02 |
| Neurological disease | 0.89 | 2.43 | 1.21–4.87 |
| Chronic renal insufficiency | 0.76 | 2.13 | 0.61–7.45 |
| Age | 0.67 | 1.96 | 0.93–4.13 |
| Malignancy | 0.44 | 1.55 | 0.69–3.48 |
| COVID-19 vaccination | 0.04 | 1.04 | 0.57–1.90 |
| COPD/asthma | −0.06 | 0.94 | 0.60–1.48 |
| Male sex | −0.13 | 0.88 | 0.50–1.56 |
| pO<sub>2</sub> | −0.29 | 0.75 | 0.23–2.41 |
| Hypertension | −0.69 | 0.50 | 0.24–1.04 |
| SpO<sub>2</sub> | −0.86 | 0.43 | 0.11–1.69 |

L2-penalized logistic regression ( $C = 1$ , class-balanced weights) fitted on all 93 patients with standardized predictors; ORs refer to one standard deviation of each predictor, including binary predictors. <sup>a</sup> Wald intervals from the observed information of the penalized, weighted fit; shown for orientation, not for formal inference

**Fig 8.**
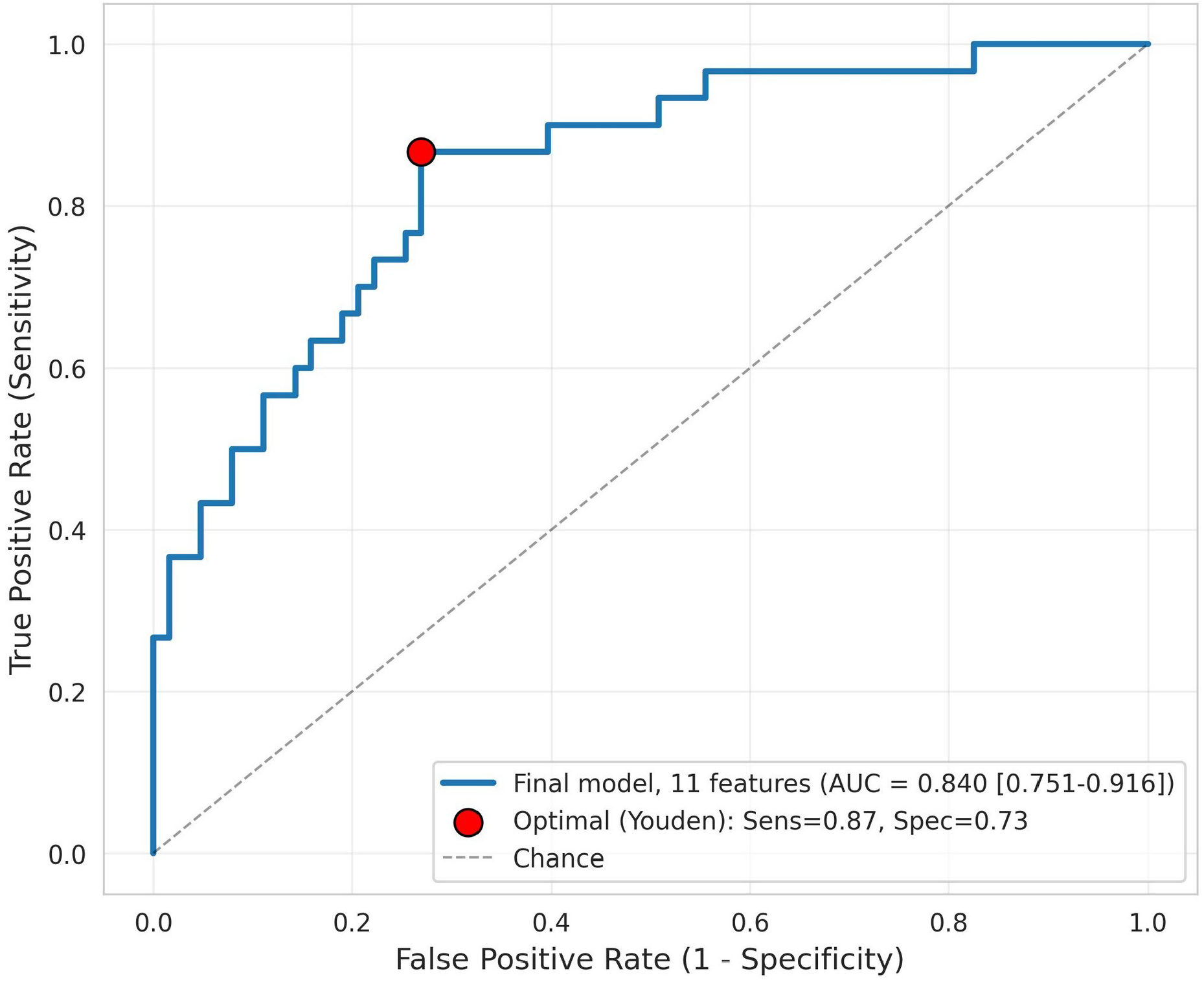
Scale D, receiver operating characteristic curve of the clinical logistic regression model (11 baseline predictors) for the composite severe outcome (high-flow oxygen therapy, mechanical ventilation or death) in the Kragujevac cohort (N = 93, 30 events), estimated by leave-one-out cross-validation. The red point marks the Youden-optimal threshold

Genotype frequencies were consistent with the European reference (rs11549465 CT/TT 12.4% vs 11.7% in 1000 Genomes; rs41508050 CT/TT 14.3%; Online Resource 1, Tables S1–S3). Mortality was 9.1% (1/11) in rs11549465 CT/TT carriers and 35.9% (28/78) in CC homozygotes (OR 0.18, exact 95% CI 0.004–1.39; p = 0.095). The clinical model discriminated similarly in both rs11549465 strata (AUC 0.82 in CC, 0.90 in CT/TT, n = 11). H3 was therefore not supported at conventional significance in this cohort.

### Cross-scale summary

Within each cohort, adding information beyond demographics improved discrimination: radiographic pneumonia in Scale A and laboratory variables in Scale C (Table 4). Across scales, the HIF1A axis appeared as convergent transcript changes, a neutrophil state associated with failed extubation, inflammatory biomarkers that dominate prediction, and a non-significant genotype association (Table 5).

**Table 4.** Discrimination by information layer within each cohort.

| Information layer | Cohort (outcome) | AUC | Within-cohort gain |
| --- | --- | --- | --- |
| Demographics and comorbidities | Mexico, N = 35,268 (ICU) | 0.61 | – |
| + radiographic pneumonia | Mexico, N = 35,268 (ICU) | 0.66 | +0.05 |
| Demographics | Brazil, N = 326 (ICU transfer) | 0.64 | – |
| + admission laboratory variables | Brazil, N = 326 (ICU transfer) | 0.79 | +0.16 |
| Clinical model, 11 predictors | Serbia, N = 93 (composite severe outcome) | 0.84 | not comparable<sup>a</sup> |
| Age and SpO<sub>2</sub> only | Serbia, N = 93 (composite severe outcome) | 0.80 | – |

Gains are computed only within a cohort. <sup>a</sup> Different population, predictors and outcome; not an increment over the Brazilian model

**Table 5.**
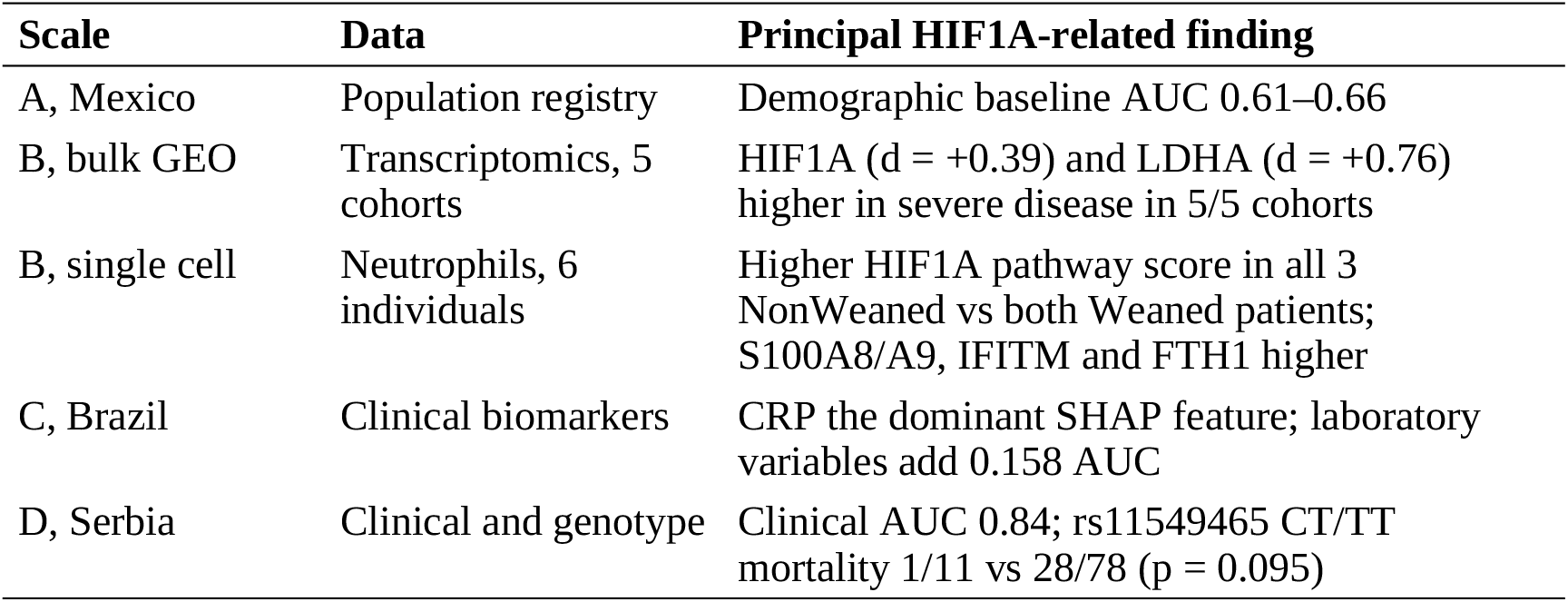
HIF1A-related findings across scales.

| Scale | Data | Principal HIF1A-related finding |
| --- | --- | --- |
| A, Mexico | Population registry | Demographic baseline AUC 0.61–0.66 |
| B, bulk GEO | Transcriptomics, 5 cohorts | HIF1A ($d = +0.39$) and LDHA ($d = +0.76$) higher in severe disease in 5/5 cohorts |
| B, single cell | Neutrophils, 6 individuals | Higher HIF1A pathway score in all 3 NonWeaned vs both Weaned patients; S100A8/A9, IFITM and FTH1 higher |
| C, Brazil | Clinical biomarkers | CRP the dominant SHAP feature; laboratory variables add 0.158 AUC |
| D, Serbia | Clinical and genotype | Clinical AUC 0.84; rs11549465 CT/TT mortality 1/11 vs 28/78 ($p = 0.095$) |

## Discussion

Four independent lines of evidence point to the same pathway. HIF1A and its glycolytic target LDHA rise with severity in every bulk cohort, a HIF1A-high, alarmin-rich neutrophil state marks patients who fail extubation, and biomarkers downstream of inflammation and organ injury carry most of the predictive signal beyond demographics. The host-genetic layer is consistent with a role for HIF1A but does not establish one.

### HIF1A as a neutrophil inflammatory axis

Under normoxia HIF1A is hydroxylated by prolyl hydroxylases and degraded through VHL [30]; hypoxia and inflammatory signals stabilize it. In myeloid cells, loss of HIF1A depletes the glycolytic energy supply and impairs aggregation, motility and invasion [8], and in neutrophils hypoxia delays apoptosis through HIF1A-dependent NF-κB activity [9]. Prolonged survival of activated neutrophils in a hypoxic lung is a plausible route from HIF1A activation to sustained tissue injury and failure to wean. The genes that separate NonWeaned from Weaned cells fit this picture. S100A8/A9 (calprotectin) is released by activated neutrophils, and high circulating calprotectin with emergency myelopoiesis distinguishes severe from mild COVID-19 [31]. FTH1 links the signature to HIF-regulated iron handling [32], and the IFITM genes reflect interferon exposure. The bulk data add the metabolic arm: LDHA had the largest and most consistent effect, in line with HIF1A-driven glycolysis in COVID-19 monocytes [10].

These are associations, not perturbations. The single-cell data come from five patients, and the group difference rests on three versus two individuals. Sequencing depth was lower in NonWeaned samples, which inflates the normalized share of highly abundant transcripts such as S100A8/A9; the direction of the alarmin signal is consistent with independent protein data [31], but its magnitude here is not reliable. Pseudobulk analysis in a larger cohort, and functional tests of HIF1A inhibition in patient neutrophils, are the next steps.

### Biomarkers as the clinical readout of the axis

CRP, a downstream product of IL-6 and NF-κB signalling that cross-talks with HIF1A [7], was the dominant predictor of ICU transfer, and urea, creatinine and LDH tracked organ dysfunction and tissue damage. In Kragujevac, LDH separated survivors from non-survivors whereas CRP did not, which suggests that admission CRP is more informative for early deterioration than for death in a cohort already selected for severe disease. Because the cohorts differ in case mix, timing and outcome definition, the AUC gains in Table 4 are within-cohort increments and should not be read as one cumulative model.

### Host genetics

rs11549465 CT/TT carriers had lower mortality, but the estimate was imprecise (exact 95% CI 0.004–1.39), the test was not significant, and power for an OR of 0.2 was about one third. Interpretation must also respect the functional data: both Pro582Ser and Ala588Thr increase HIF1A transactivation in reporter assays [12], so a protective association would not follow from a simple model in which less HIF1A activity is beneficial. Context-dependent effects, such as earlier adaptive responses to hypoxia, are possible but untested. These variants are absent from standard COVID-19 genotyping panels [13], and a pooled multi-locus analysis with the interferon-lambda and ACE2/TMPRSS2 variants previously studied at this center [14–16], with sex-stratified effects, is the appropriate design for testing them.

### Hypertension

Hypertension had a negative coefficient in the Kragujevac model. Chronic ACE inhibitor or angiotensin receptor blocker therapy is a possible explanation: discontinuing these drugs did not improve outcome in a randomized trial [33], and prior use was not associated with severe disease in a large case-control study [34]. Outpatient medication was not recorded uniformly in our cohort, the coefficient comes from a penalized model, and removing hypertension changed the AUC by only 0.026, so we treat this as a hypothesis.

### Limitations

First, the scales differ in population, outcome (ICU admission, death, composite severe outcome) and timing, so their AUCs are not directly comparable. Second, the Kragujevac cohort is small (about 2.7 events per predictor); imputation and standardization were estimated before cross-validation, and coefficients from the penalized, class-weighted model are not suitable for formal inference. Third, the single-cell dataset includes five patients and one healthy donor, with depth differences between groups, so cell-level p values overstate certainty. Fourth, patients were treated during the first pandemic wave, before routine use of remdesivir, tocilizumab or baricitinib. Fifth, the Scale C model did not transfer to Kragujevac, reflecting different variables and outcomes. Sixth, sex was included as a covariate, but sex-stratified genotype effects could not be estimated reliably at this sample size; our earlier work at this center found female-specific associations for other loci [15, 16], so future studies should be powered for sex-stratified analysis.

### Conclusions

Demographics and comorbidities set a baseline for predicting severe COVID-19 that inflammatory and organ-dysfunction biomarkers exceed within each cohort. HIF1A pathway activation is reproducible across five transcriptomic cohorts and localizes to an alarmin-rich neutrophil state in patients who fail extubation. The HIF1A ODD-domain variant rs11549465 showed a non-significant association with lower mortality that warrants testing in larger, sex-balanced cohorts. Together, the data support the HIF1A axis as a target for mechanistic studies of neutrophil-driven inflammation in severe viral pneumonia.

## Supporting information

Online Resource 1. Supplementary methods and tables

Online Resource 2. Supplementary figures S8-S20

Online Resource 3. TRIPOD checklist

## Data Availability

Public datasets are available from their providers: the Mexican Directorate General of Epidemiology open COVID-19 data (https://www.gob.mx/salud/documentos/datos-abiertos-152127); NCBI GEO accessions GSE152075, GSE157103, GSE171110, GSE212861, GSE300696 and GSE234904; and the Sirio-Libanes COVID-19 dataset on Kaggle. All analysis code, derived numerical results and trained models are available at https://github.com/zivanovicmkg/hif1a-covid-multiscale and archived on Zenodo (https://doi.org/10.5281/zenodo.20594498). Patient-level data of the Kragujevac cohort are not publicly available under the ethics approval; de-identified data are available from the corresponding author on reasonable request, subject to review and a data-access agreement.

## Declarations

### Ethics approval

All procedures involving human participants were performed in accordance with the 1964 Declaration of Helsinki and its later amendments. The study “Investigation of the humoral response and of phenotypic and functional features of peripheral blood cells in patients with COVID-19” was approved by the Ethics Committee of the Clinical Center Kragujevac (No. 01/20-498, 5 May 2020). Analyses of public datasets (Scales A–C) used de-identified data released for secondary research and did not require additional approval.

### Consent to participate

Written informed consent was obtained from all individual participants of the Kragujevac cohort.

### Consent to publish

Not applicable. The article contains no individual-level identifying information or images.

### Data availability

Public datasets are available from their providers: Mexican Directorate General of Epidemiology open data (https://www.gob.mx/salud/documentos/datos-abiertos-152127); NCBI GEO accessions GSE152075, GSE157103, GSE171110, GSE212861, GSE300696 and GSE234904; and the Sírio-Libanês COVID-19 dataset on Kaggle. Derived numerical results are archived on Zenodo (https://doi.org/10.5281/zenodo.20594498) under CC BY 4.0. Patient-level data of the Kragujevac cohort are not publicly available under the ethics approval; de-identified data are available from the corresponding author on reasonable request, subject to review and a data-access agreement.

### Code availability

All analysis code is available at https://github.com/zivanovicmkg/hif1a-covid-multiscale and archived on Zenodo (https://doi.org/10.5281/zenodo.20594498) under the MIT License.

### Author contributions

Conceptualization: Marko N. Živanović, Ivana Raković, Biljana Popovska Jovičić; Methodology: Marko N. Živanović; Software: Marko N. Živanović; Formal analysis: Marko N. Živanović, Miloš Z. Milosavljević; Investigation (patient recruitment and clinical data): Ivana Raković, Biljana Popovska Jovičić, Miloš Z. Milosavljević; Investigation (DNA extraction and genotyping): Ana Mirić, Nevena Milivojević Dimitrijević, Jelena Košarić, Katarina Virijević, Marija Branković, Tamara Mladenović; Resources: Marko N. Živanović (computational resources), Ivana Raković, Biljana Popovska Jovičić (clinical resources); Data curation: Marko N. Živanović; Visualization: Marko N. Živanović; Project administration: Marko N. Živanović; Ivana Raković, Biljana Popovska Jovičić (ethics approval); Supervision: Marko N. Živanović; Funding acquisition: Marko N. Živanović, Nenad Filipović; Writing – original draft: Marko N. Živanović, Ivana Raković, Biljana Popovska Jovičić, Ana Mirić, Nevena Milivojević Dimitrijević, Jelena Košarić, Katarina Virijević, Marija Branković, Tamara Mladenović; Writing – review and editing: all authors. All authors read and approved the final manuscript.

### Use of generative AI

Claude Code (Anthropic) was used to draft and refactor analysis scripts (data preprocessing, machine-learning pipelines and statistical routines). Claude (Anthropic) was used for language editing, restructuring the manuscript to the journal’s format and checking consistency between the manuscript and the code repository. AI tools were not used to generate data, results, figures or citations. The authors reviewed and verified all code and text and take full responsibility for the content.

### Supplementary Information

**Online Resource 1** Supplementary methods (filter cascades, Harmony parameters, genotyping and Hardy–Weinberg tables), supplementary tables S4–S8, extended biological context of the HIF1A pathway and the literature analysis (PDF)

**Online Resource 2** Supplementary figures S8–S20 (PDF)

**Online Resource 3** TRIPOD checklist (PDF)

