## Supplementary material for "Multi-scale analysis of the HIF1A hypoxia–inflammation axis in severe COVID-19": Online Resource 1. Supplementary methods and tables

### S1. Additional methodology details

#### S1.1 Scale A — Mexico CDC filter cascade

The initial Mexican CDC dataset (2020–2022 cumulative) contained 263.214 recorded cases of COVID-19 [1]. The primary filter on PCR-confirmed cases retained 103.452 cases. The secondary filter on hospitalized cases retained 62.184 cases. A tertiary filter on patients with complete data for all predictor variables yielded a final analytic cohort of N=35.268. For all models, data were split 70/30 stratified by outcome (ICU or mortality). ML hyperparameters: LR with L2 regularization C=1.0; RF with 300 trees and max_depth=15; XGBoost with learning_rate=0.1, max_depth=6, subsample=0.8. Class imbalance is addressed by class_weight='balanced' for LR/RF or scale_pos_weight for XGBoost.

#### S1.2 Scale B — bulk meta-analysis details

The five GEO cohorts were downloaded programmatically from the GEO DataSets API and their GSE identifiers. Expression matrices in raw scales (TPM, FPKM or raw counts in different cohorts) are not directly comparable, which is the reason for per-cohort z-score standardization. The gene panel was selected based on the HIF1A literature and pre-defined before analysis: HIF1A (master regulator), VEGFA (vascular), SLC2A1/LDHA/PDK1 (glycolytic axis), IL6/TNF/CXCL8 (inflammatory cytokines), STAT3 (cross-talk with NF-κB) and MMP9 (extracellular matrix remodeling).

#### S1.3 Scale B — scRNA-seq Harmony parameters

Harmony integration was applied with the following parameters: basis='X_pca' (50 PCs), key='sample_id', max_iter_harmony=10 (converged for 4 iterations), epsilon_cluster=1e-5, epsilon_harmony=0.0001, sigma=0.1, theta=2 (moderate correction strength). Post-Harmony kNN: n_neighbors=15, n_pcs=30. Leiden: resolution 0.5, random_state=42. The HIF1A pathway score was calculated by the scanpy score_genes function with n_bins=25 and ctrl_size=50 to define the background gene set.

#### S1.4 Scale C — Sírio-Libanês filtering and outcome

The original Sirio-Libanes Kaggle release (Oct 2020) [2] contains 1,925 serial observations from 385 patients with 5-time windows: 0–2h, 2–4h, 4–6h, 6–12h, 12+h post-admission. For the predictive framework "who will transfer to the ICU in the next 4–12h", only variables from the 0–4h window were used, and the outcome was defined as ICU admission in any of the later windows (4+h). Patients who were already in the ICU at admission were excluded to avoid data leakage (385 → 326). Features with <50% non-missing coverage are excluded. Final N=326, 30 variables, event rate (ICU later) = 41.7%.

#### S1.5 Scale D — DNA extraction and HIF1A SNP genotyping (MIQE-aligned)

Genotyping of HIF1A polymorphisms was performed in accordance with laboratory protocols aligned with the MIQE guidelines [3] and Good Laboratory Practice (GLP) propositions for PCR detection and sample-centric laboratory handling. Because the present application is allelic discrimination for genotyping rather than quantitative gene expression, MIQE items relating to reverse transcription, Cq-based normalization, and ΔΔCq analysis are not applicable to this experimental design; remaining MIQE principles regarding sample handling, nucleic acid quality, target identification, oligonucleotide documentation, reaction conditions, quality control, and data analysis were fully applied.

**Sample handling**. Peripheral venous blood was collected in K2-EDTA vacuum tubes (BD Vacutainer, 4 mL) on the day of patient admission, prior to initiation of corticosteroid or antiviral therapy. Samples were transported on ice to the molecular biology laboratory and processed within 4 hours of collection. Aliquots not processed immediately were stored at +4 °C and processed within 24 hours, or frozen at −80 °C for longer storage.

**DNA extraction**. Genomic DNA was extracted from 200 μL of whole blood using the PureLink™ Genomic DNA Mini Kit (Cat. No. K182002, Invitrogen, Thermo Fisher Scientific), following the manufacturer's spin-column protocol. Briefly, samples were lysed with proteinase K and Lysis Buffer at 55 °C for 10 minutes; ethanol precipitation was followed by two wash steps with Wash Buffer 1 and Wash Buffer 2; DNA was eluted in 50 μL of Elution Buffer. Extracted DNA was used immediately for downstream genotyping or stored at −20 °C.

**DNA quantification**. DNA concentration and purity were determined by UV spectrophotometry on a NanoDrop instrument (Thermo Fisher Scientific) at 260 nm. The $A_{260}/A_{280}$ absorbance ratio was used as the primary quality criterion (acceptable range 1.75–2.00, indicating absence of protein, phenol, and RNA contamination). Samples failing this threshold were re-extracted from archived blood aliquots.

**Target gene and SNP information**. Two HIF1A single-nucleotide polymorphisms located within the oxygen-dependent degradation (ODD) domain — a region critical for prolyl hydroxylase domain (PHD)-mediated protein degradation under normoxic conditions were genotyped. Complete target gene information including chromosomal positions, allele designations, amino acid changes, and TaqMan Assay IDs is summarized in Table S1.

**Genotyping assays**. Allelic discrimination was performed using pre-validated TaqMan™ SNP Genotyping Assays (Applied Biosystems, Thermo Fisher Scientific): rs11549465 — Assay ID C__25473074_10 (Lot P230911-011B06); rs41508050 — Assay ID C__86499352_10 (Lot P230911-011B07). Each assay contains two allele-specific TaqMan minor-groove binder (MGB) probes labeled with VIC (allele C) and FAM (allele T) fluorophores, plus an unlabeled primer pair, supplied as a single 40× reagent mixture. Probe sequences are proprietary to the manufacturer; functional validation data are available from the supplier (https://www.thermofisher.com).

**qPCR reaction setup**. Allelic discrimination reactions were performed in a final volume of 20 μL containing 10 μL of TaqMan™ Genotyping Master Mix (2×; Cat. No. 4371355, Lot 2696189; Applied Biosystems), 1 μL of 20× SNP Genotyping Assay mix, and 9 μL of template DNA solution (approximately 10 ng total DNA per reaction, diluted in nuclease-free water). Reactions were assembled in Mic qPCR optical tubes pre-loaded with high-viscosity silicon oil to prevent evaporation, briefly centrifuged, and run on the instrument. Dual-filter pipette tips were used throughout all sample handling to prevent cross-contamination.

**Thermal cycling conditions**. Reactions were run on the Mic qPCR Cycler (Biomolecular Systems, Yatala, Australia) controlled by BMS Workbench v2.0 software. Cycling protocol: AmpliTaq Gold polymerase activation at 95 °C for 10 minutes, followed by 40 cycles of denaturation at 95 °C for 15 seconds and annealing/extension at 60 °C for 60 seconds. Fluorescence acquisition (VIC and FAM channels) occurred at the end of each annealing/extension step. Endpoint fluorescence values were used for allelic discrimination calling.

**Quality controls**. Every plate included at least one no-template control (NTC) with nuclease-free water replacing the DNA template; all NTCs showed no amplification (Cq > 40). All patient samples were processed in technical duplicate to assess inter-run reproducibility. Duplicate concordance was excellent: 95.2% for rs11549465 (40/42 concordant pairs; 2 discordant) and 100% for rs41508050 (41/41 concordant pairs), with overall combined concordance of 97.6% (81/83). The two discordant samples for rs11549465 (patients 84 and 119) were reviewed manually on the allelic discrimination scatter plot; both showed intermediate fluorescence signatures consistent with heterozygous (CT) genotype calls in at least one replicate, supporting a final assignment of CT carrier status. Complete duplicate concordance statistics are provided in Table S3.

**Data analysis**. Genotype calls were obtained from endpoint VIC and FAM fluorescence values using BMS Workbench v2.0 allelic discrimination analysis, with a fluorescence threshold of 1.5 (starting at cycle 1). Allelic discrimination scatter plots were manually reviewed for every plate by an independent operator. Samples with ambiguous clustering were re-assayed in duplicate; final genotypes were assigned only when concordant calls were obtained in at least two of three assays. Samples for which a confident call could not be obtained after two repeat assays were excluded from genetic analyses, yielding a final analytical N = 89 for rs11549465 and N = 77 for rs41508050 from the 93-patient cohort.

**Reference population validation**. Observed minor allele frequencies in our cohort were compared with the European (EUR) reference from the 1000 Genomes Project Phase 3: rs11549465 MAF = 0.062 (cohort) vs. 0.08 (1000G-EUR); rs41508050 MAF = 0.092 (cohort) vs. 0.09 (1000G-EUR). Observed frequencies are consistent with the European reference, supporting absence of selection bias or systematic genotyping artifact.

**Hardy–Weinberg equilibrium**. HWE was tested by chi-square goodness-of-fit (1 degree of freedom for a biallelic SNP) on observed three-genotype counts from technical replicates. No significant deviation from HWE was detected for either polymorphism (rs11549465: $\chi^2 = 0.32$, $p = 0.575$; rs41508050: $\chi^2 = 0.61$, $p = 0.434$; Table S2). Sanger sequencing validation was not performed; this limitation is acknowledged in the main manuscript.

Table S1 provides complete target gene information for the two HIF1A single-nucleotide polymorphisms genotyped in the Kragujevac clinical-genetic cohort. Both polymorphisms are located in exon 12 of HIF1A, within the oxygen-dependent degradation (ODD) domain.

| Parameter | rs11549465 | rs41508050 |
|---|---|---|
| Gene locus | HIF1A | HIF1A |
| Chromosome | 14q23.2 | 14q23.2 |
| Genomic position (GRCh38) | 14:61740839 | 14:61738090 |
| Reference sequence | NC_000014.9 | NC_000014.9 |
| cDNA change | c.1744C>T | c.1762G>A |
| Amino acid change | p.Pro582Ser | p.Ala588Thr |
| Functional domain | ODD domain | ODD domain |
| TaqMan Assay ID | C__25473074_10 | C__86499352_10 |
| Assay lot number | P230911-011B06 | P230911-011B07 |
| Alleles (major/minor) | C / T | G / A |
| Fluorophore labels | VIC (C) / FAM (T) | VIC (C) / FAM (T) |
| MAF (EUR, 1000G Phase 3) | 0.08 | 0.09 |
| MAF (Kragujevac cohort) | 0.062 | 0.092 |
| CT/TT carriers, n (% of N=89/77) | 11 (12.4%) | 11 (14.3%) |

*Table S1 — HIF1A SNP target information for TaqMan allelic discrimination. ODD = oxygen-dependent degradation domain; MAF = minor allele frequency; EUR = European reference population; 1000G = 1000 Genomes Project Phase 3.*

Hardy–Weinberg equilibrium (HWE) was assessed for both polymorphisms using observed three-genotype counts derived from technical replicates of TaqMan allelic discrimination assays. Expected genotype counts were calculated from observed allele frequencies under the assumption $p^2 + 2pq + q^2 = 1$, and deviation was tested by chi-square goodness-of-fit (1 degree of freedom for a biallelic SNP). Both polymorphisms satisfied HWE ($p > 0.05$), supporting the integrity of the genotyping protocol and absence of population stratification or selection bias.

| SNP | N | Observed (CC / CT / TT) | MAF (q) | Expected (CC / CT / TT) | $\chi^2$ | p-value |
|---|---|---|---|---|---|---|
| rs11549465 | 73 | 64 / 9 / 0 | 0.062 | 64.3 / 8.4 / 0.3 | 0.32 | 0.575 |
| rs41508050 | 60 | 49 / 11 / 0 | 0.092 | 49.5 / 10.0 / 0.5 | 0.61 | 0.434 |

*Table S2 — Hardy–Weinberg equilibrium assessment for both HIF1A polymorphisms. N refers to the number of patients with concordant duplicate calls confirmed from the available .micrun raw data; this differs slightly from the N in the analytical dataset (89 for rs11549465 and 77 for rs41508050) which includes additional patients genotyped on alternative runs not available in the current data export. Both polymorphisms show no significant deviation from HWE ($p > 0.05$). MAF = minor allele frequency.*

All patient samples were processed in technical duplicate to assess inter-run reproducibility of TaqMan allelic discrimination calls. Concordance was evaluated patient-wise: a concordant patient is one with identical genotype calls in both replicate reactions. Two discordant calls were observed for rs11549465 (patients 84 and 119), both of which showed intermediate fluorescence signatures on the allelic discrimination scatter plot, with at least one replicate calling Heterozygous (CT). Final genotype assignments for discordant patients were made by manual review of the scatter plot and majority-rule from the duplicate calls.

| SNP | Patients with | Concordant pairs | Discordant pairs | Concordance rate |
|---|---|---|---|---|

| | duplicates | | | |
|---|---|---|---|---|
| rs11549465 | 42 | 40 | 2 | 95.2% |
| rs41508050 | 41 | 41 | 0 | 100.0% |
| Combined | 83 | 81 | 2 | 97.6% |

*Table S3 — Inter-run duplicate concordance for HIF1A allelic discrimination. Overall concordance was 97.6% across both SNPs, exceeding the MIQE-recommended threshold of 95% for genotyping reproducibility. Discordant calls (n=2) were manually reviewed and resolved by scatter plot inspection.*

### S2. Additional figures

#### S2.1 Scale B supplementary figures

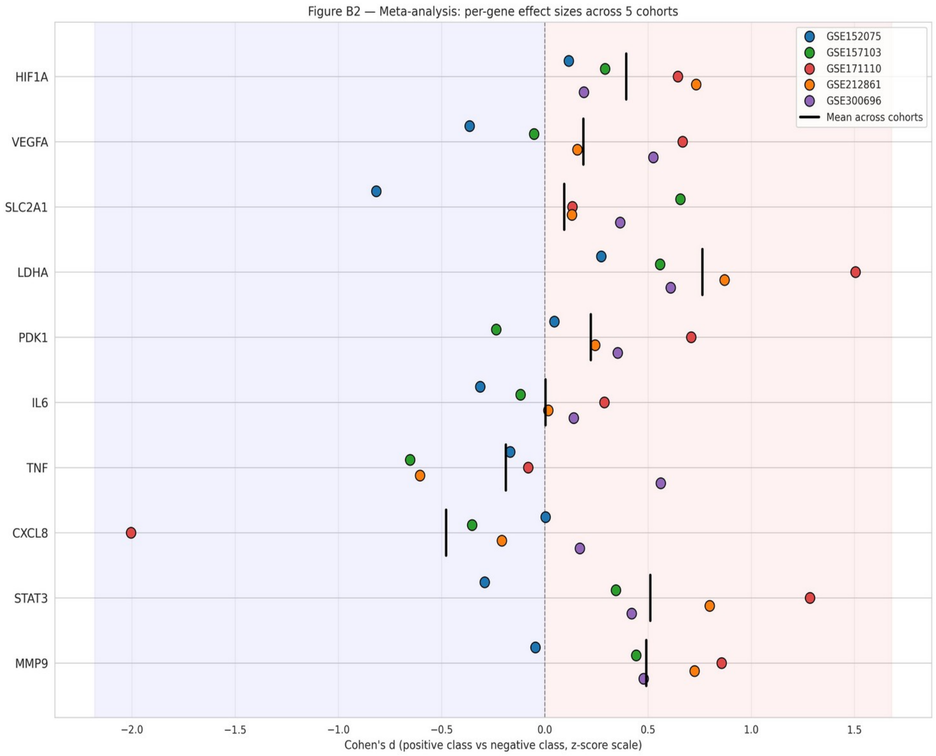


*Figure S1 — Scale B bulk, forest plot of Cohen's d effect-size meta-analyses by gene and cohort. Presented as a complement to Fig. 2 of the main text (heatmap). Each line = gene; single colored dots = effect in each of 5 cohorts; vertical black lines = mean d across cohorts. HIF1A, LDHA, STAT3 and MMP9 show consistently positive mean values across cohorts.*

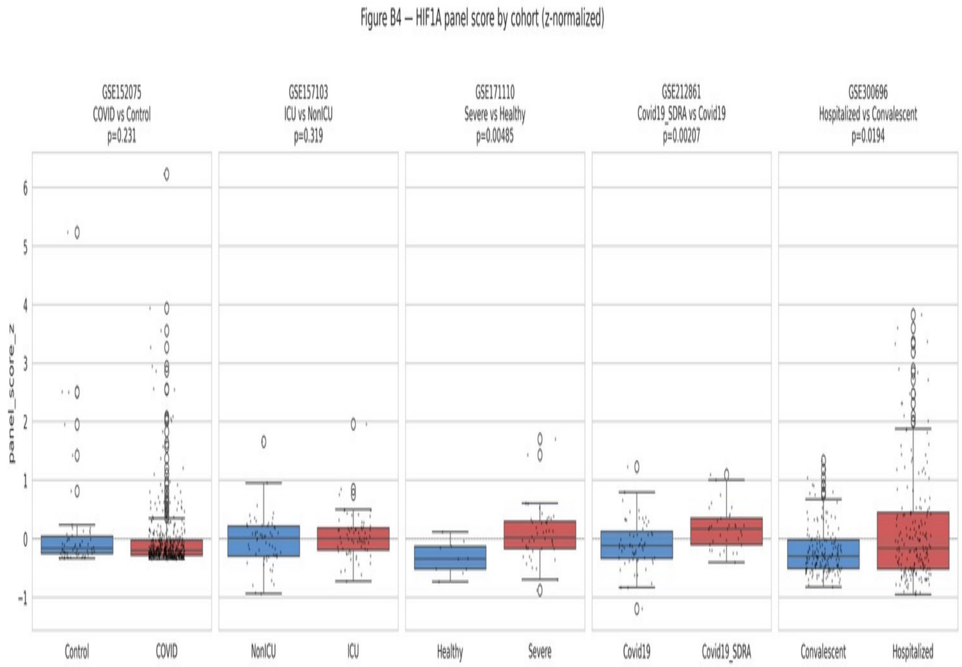


*Figure S2 — Scale B bulk, boxplot of panel_score_z distributions (simple arithmetic mean of 10 standardized gene values) by cohort and class. Mann-Whitney U test p-values are shown. GSE171110 (Severe vs Healthy) and GSE212861 (Covid19_SDRA vs Covid19) show significant differences, while GSE152075 (Covid vs Control contaminated by healthcare workers as "controls") does not reach significance. This illustrates why multivariate panel LR is superior to simple averaging.*

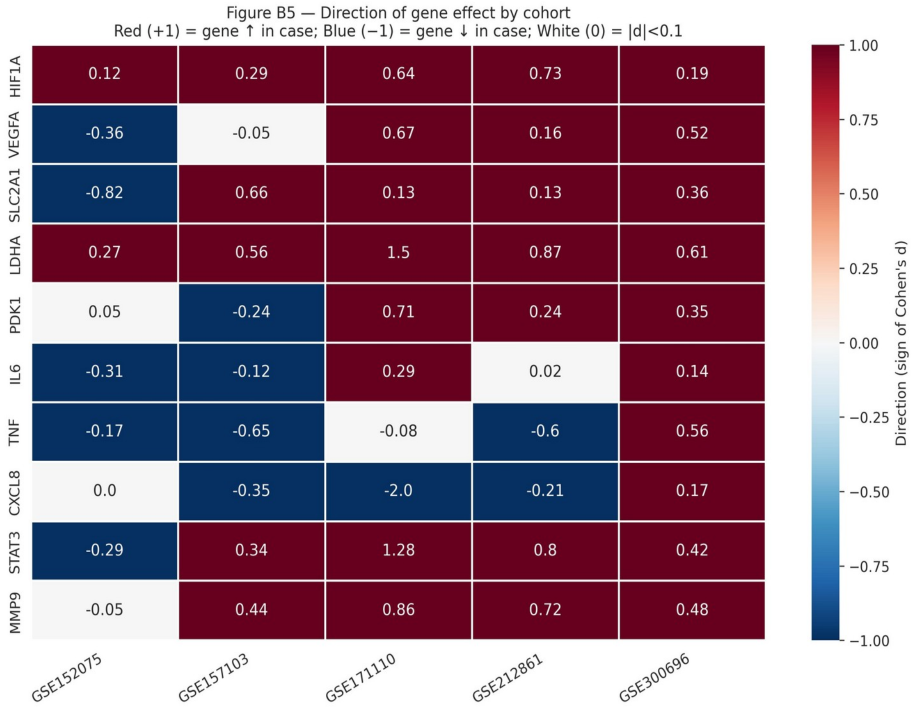


*Figure S3 — Scale B bulk, alternative visualization of consistency of direction of effects by gene and cohort. This is a supplementary figure that is replaced by the main display with a heatmap (Fig. 2 of the main text). Included for completeness of material.*

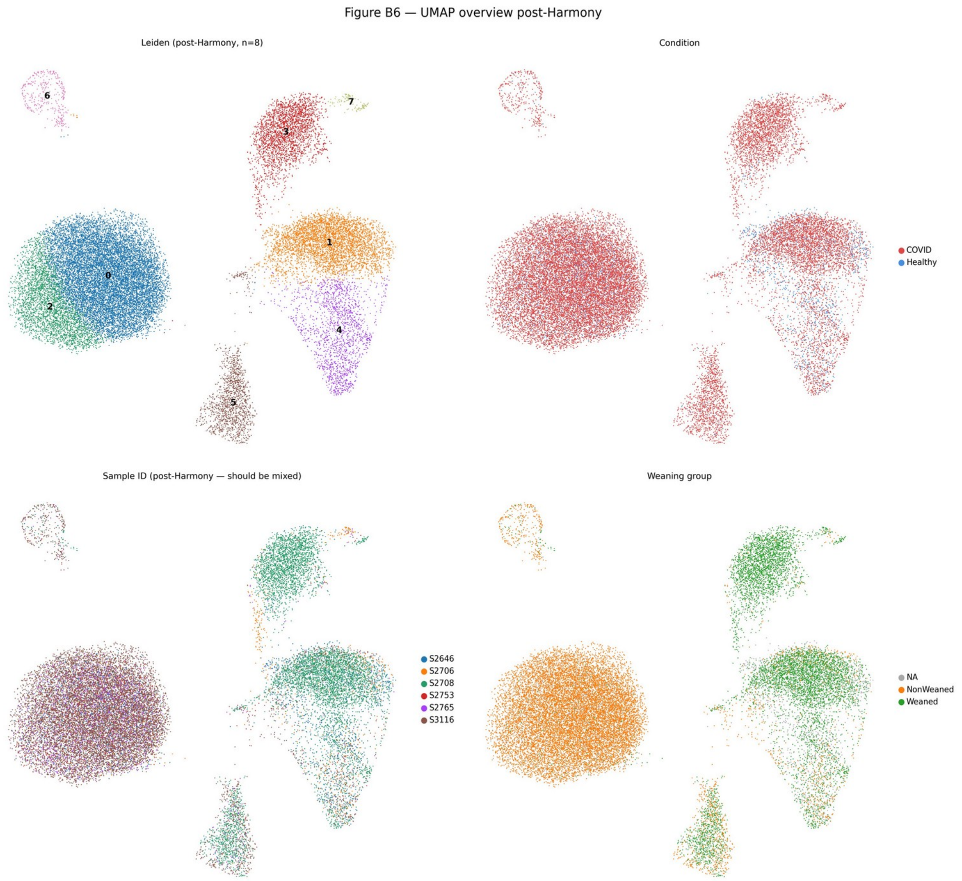


*Figure S4 — Scale B scRNA-seq, post-Harmony UMAP 4-panel overview: clusters, condition (COVID vs Healthy), sample (post-Harmony shuffling), extubation group. This is a supplement to Online Resource 2, Fig. S12 (before/after Harmony comparison) and shows the final biologically-driven result.*

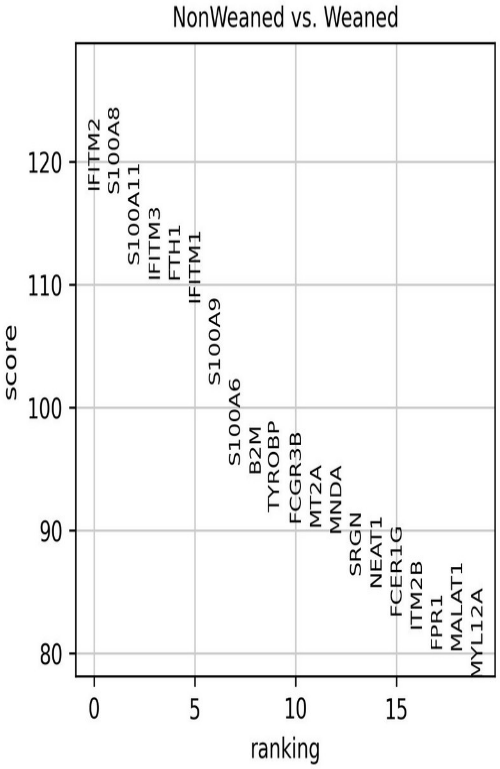


*Figure S5 — Scale B scRNA-seq, bar plot of top 20 DEG genes ranked by the Wilcoxon rank-sum test statistic. Alternative display of data from Fig. 5 of the main text (heatmap). All genes have an adjusted p < 0.001. Top ranked: IFITM2, S100A8, S100A11, IFITM3, FTH1, IFITM1, S100A9.*

#### S2.2 Scale C supplementary figures

The main C figure scales are C1 (ROC), C4 (calibration), C5 (SHAP + DCA), C6 (top features + incremental). There are no additional figures for Scale C in this material; all relevant analyzes are included in the main manuscript.

#### S2.3 Scale D supplementary figures

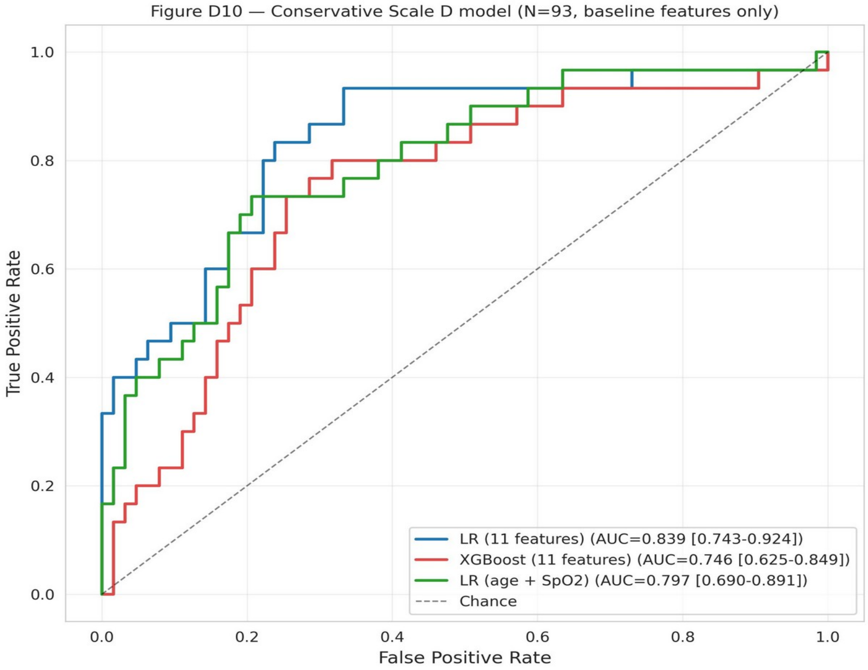


*Figure S6 — Scale D, conservative LR model ROC curve (alternative to the final model shown in Fig. 8 of the main text). This model retains comorbidities_bin (aggregated comorbidity variable) as an additional variable; performance is comparable to the final model but comorbidities_bin is strongly correlated with HTA, creating multicollinearity. The final model (Option B) removes comorbidities_bin for interpretability.*

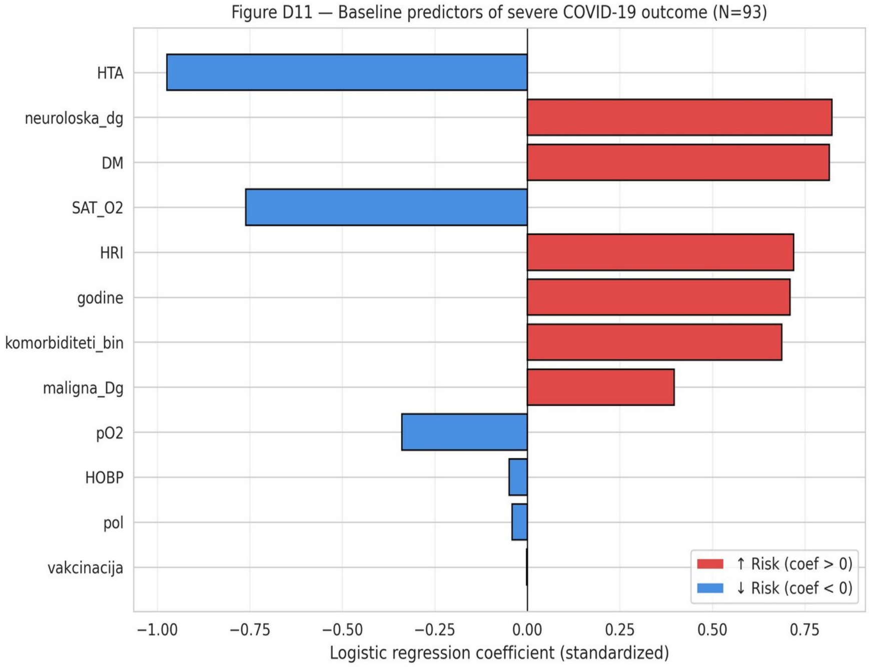


*Figure S7 — Scale D, LR coefficients of the conservative model (with comorbidities_bin). Supplement to Online Resource 2, Fig. S18. The same variables are identified as dominant predictors, but the HTA paradox is somewhat mitigated due to the correlation with comorbidities_bin.*

### S3. Additional tables

#### S3.1 Therapies applied in the Kragujevac cohort

| Therapy | Total (N=93) | Survived (N=64) | Deceased (N=29) | Note |
|---|---|---|---|---|
| Antibiotic | 93 (100%) | 64 (100%) | 29 (100%) | routine use in protocol |
| Corticosteroids | 92 (98,9%) | 63 (98,4%) | 29 (100%) | according to availability and protocol |
| Oxygen therapy | 93 (100%) | 64 (100%) | 29 (100%) | all patients at some level of $O_2$ |
| Favipiravir | 38 (40,9%) | 29 (45,3%) | 9 (31,0%) | Fisher p=0,256 |
| Remdesivir | 17 (18,3%) | 11 (17,2%) | 6 (20,7%) | Fisher p=0,774 |
| Tocilizumab | 0 (0%) | 0 (0%) | 0 (0%) | was not available during the period |
| Mechanical ventilation | 9 (9,7%) | 0 (0%) | 9 (31,0%) | Fisher p<0,001 |
| High-flow (HFV) | 19 (20,4%) | 1 (1,6%) | 18 (62,1%) | Fisher p<0,001 |

*Table S4 — Applied therapies in the Kragujevac cohort. Antibiotics, corticosteroids and oxygen therapy were administered to (almost) all patients as part of the routine protocol. Significant differences between survivors and deceased are in the use of mechanical ventilation and HFV, which is expected given the escalation of therapy with the severity of the condition.*

#### S3.2 Scale B — per-cohort AUC details

| Cohort | Comparison | N (positive) | N (negative ) | LR CV AUC | panel_score AUC |
|---|---|---|---|---|---|
| GSE152075 | COVID vs Control | 430 | 54 | 0,753 | 0,450 |
| GSE157103 | ICU vs NonICU | 65 | 58 | 0,718 | 0,552 |
| GSE171110 | Severe vs Healthy | 44 | 10 | 0,989 | 0,789 |
| GSE212861 | Covid19_SDRA vs Covid19 | 33 | 60 | 0,779 | 0,694 |
| GSE300696 | Hospitalized vs Convalescent | 188 | 158 | 0,841 | 0,573 |

*Table S5 — Scale B bulk, per-cohort AUC performance. LR CV AUC: multivariate logistic regression model with 5-fold cross-validation. panel_score AUC: simple arithmetic mean of standardized gene values.*

#### S3.3 Scale B — an effect-size meta-analysis

| Gene | GSE152075 | GSE157103 | GSE171110 | GSE212861 | GSE300696 | mean d |
|---|---|---|---|---|---|---|
| *HIF1A* | +0,12 | +0,29 | +0,64 | +0,73 | +0,19 | +0,39 |
| *VEGFA* | −0,36 | −0,05 | +0,67 | +0,16 | +0,52 | +0,19 |
| *SLC2A1* | −0,82 | +0,66 | +0,13 | +0,13 | +0,36 | +0,09 |
| *LDHA* | +0,27 | +0,56 | +1,50 | +0,87 | +0,61 | +0,76 |
| *PDK1* | +0,05 | −0,24 | +0,71 | +0,24 | +0,35 | +0,22 |
| *IL6* | −0,31 | −0,12 | +0,29 | +0,02 | +0,14 | +0,00 |
| *TNF* | −0,17 | −0,65 | −0,08 | −0,60 | +0,56 | −0,19 |
| *CXCL8* | +0,00 | −0,35 | −2,00 | −0,21 | +0,17 | −0,48 |
| *STAT3* | −0,29 | +0,34 | +1,28 | +0,80 | +0,42 | +0,51 |
| *MMP9* | −0,05 | +0,44 | +0,86 | +0,72 | +0,48 | +0,49 |

*Table S6 — Scale B bulk, complete matrix of Cohen's d effect-sizes for all 10 panel genes across all 5 cohorts, with means. HIF1A and LDHA are 5/5 positive (d > 0 in all cohorts); STAT3, MMP9, PDK1 are 4/5 positive; TNF and CXCL8 have heterogeneous/reversed direction.*

#### S3.4 Scale C — model performance with bootstrap CI

| Model | CV AUC (mean ± SD) | Test AUC | 95% CI | Brier | Avg Precision |
|---|---|---|---|---|---|
| Logistic regression | 0,699 ± 0,058 | 0,695 | 0,560–0,818 | 0,239 | 0,631 |
| Random Forest | 0,763 ± 0,097 | 0,754 | 0,624–0,868 | 0,200 | 0,663 |
| XGBoost | 0,761 ± 0,103 | 0,780 | 0,650–0,888 | 0,209 | 0,736 |
| LightGBM | 0,745 ± 0,102 | 0,779 | 0,665–0,881 | 0,226 | 0,725 |
| XGBoost (Platt- calibrated) | — | 0,794 | 0,686–0,887 | 0,194 | 0,707 |

*Table S7 — Scale C, detailed model performance with bootstrap 95% CI. Calibrated XGBoost achieves the best combination of AUC and Brier value.*

#### S3.5 Scale C — top 10 SHAP variables

| Rank | Variable | mean \|SHAP\| | Biological interpretation |
|---|---|---|---|
| 1 | CRP | 0,77 | Systemic inflammation — downstream of HIF1A-NF-κB |
| 2 | Diastolic BP | 0,54 | Hemodynamic — CV compensation |
| 3 | Urea | 0,50 | Renal dysfunction — HIF1A renal target |

| | | | |
|---|---|---|---|
| 4 | Age percentile | 0,49 | Demographic — age-related fragility |
| 5 | Temperature | 0,46 | Inflammatory pyrexia |
| 6 | Creatinine | 0,45 | Renal dysfunction |
| 7 | Sodium | 0,42 | Volume/fluid status |
| 8 | Heart rate | 0,37 | Hemodynamic — compensatory tachycardia |
| 9 | AST | 0,34 | Glycolytic / hepatic stress — LDHA co-elevated |
| 10 | Lymphocytes | 0,29 | Immune dysregulation — severe COVID lymphopenia |

*Table S8 — Scale C, top 10 variables by SHAP values in the calibrated XGBoost model with biological interpretations. CRP dominates with ~1.5× higher mean |SHAP| from another variable. Three biological groups: systemic inflammation, organ dysfunction, demographic-hemodynamic status.*

### S4. The HIF1A pathway—an expanded biological context

#### S4.1 Mechanism of HIF1A stabilization

Under normoxic conditions, the HIF1A protein is subject to continuous degradation through prolyl hydroxylation of residues Pro402 and Pro564 by prolyl-hydroxylase domain proteins (PHD1, PHD2, PHD3) [4]. Hydroxylated HIF1A is recognized by von Hippel-Lindau (VHL) E3 ubiquitin ligase and ubiquitinated for degradation [5]. Under hypoxic conditions or conditions that disrupt PHD/VHL activity (eg, inflammatory conditions with elevated ROS, altered Krebs cycle metabolites such as succinate and fumarate), HIF1A is stabilized, translocated to the nucleus, dimerizes with HIF1B (ARNT) and binds HRE (hypoxia response element) sequences in target gene promoters [6].

#### S4.2 Target genes in the panel

The panel of 10 genes we used includes direct (HRE-driven) and indirect HIF1A targets:

- LDHA: lactate dehydrogenase A, converts pyruvate to lactate. Direct HRE-driven target. Upregulation allows tumors/hypoxic cells to resume glycolysis in the absence of oxidative phosphorylation.
- SLC2A1 (GLUT1): glucose transporter, direct HRE target. Increased glucose import enhances glycolytic flux.
- PDK1: pyruvate dehydrogenase kinase 1, inhibits PDH and diverts pyruvate to lactate instead of the Krebs cycle. HIF1A target.
- VEGFA: vascular endothelial growth factor A, inducer of angiogenesis. HRE-driven.
- HIF1A alone: an autoregulatory loop.
- STAT3: non-direct HIF1A target, but has cross-talk through IL-6/JAK signaling that affects HIF1A stability.
- NF-κB axis (IL6, TNF, CXCL8): inflammatory cytokines whose expression cross-talks with HIF1A through common kinases.

- MMP9: matrix-metalloproteinase 9, neutrophil effector, regulated by both HIF1A and NF-κB.

##### S4.3 rs11549465 functional characterization

rs11549465 (c.1744C>T, p.Pro582Ser) is located in exon 12 of the HIF1A gene and results in the replacement of proline by serine at position 582 of the protein. This position is located in the ODD (oxygen-dependent degradation) domain, a region critical for PHD-mediated degradation. In reporter assays, both Pro582Ser and Ala588Thr showed higher transcriptional activity than the wild-type protein under normoxic and hypoxic conditions (Tanimoto et al., Carcinogenesis 2003;24:1779–83; https://doi.org/10.1093/carcin/bgg132). In the Kragujevac cohort, mortality was 1/11 in rs11549465 CT/TT carriers and 28/78 in CC homozygotes (conditional maximum-likelihood OR 0.18, exact 95% CI 0.004–1.39; Fisher's exact p = 0.095). Given the wide interval and the direction of the published functional effect, this association is not interpreted mechanistically; it requires replication in a larger cohort.

#### Supplementary Section S5 — COVID-19 Literature Mining Analysis

This section reports an automated bibliometric analysis of the COVID-19 literature (2019/01/01–2026/06/01) performed to contextualize the present multi-scale HIF1A study. Five PubMed query groups (G1–G5) were executed via NCBI Entrez and ranked by OpenAlex citation count. Full pipeline details, complete reference lists, and raw data are archived in the project repository (cited in the main text).

##### S5.1 Search strategy

Literature retrieval used NCBI Entrez (Biopython) with an institutional API key. Five parallel query groups were defined and executed with date range 2019/01/01–2026/06/01: G1 (COVID-19; MeSH OR title/abstract), G2 (COVID-19 + AI/ML), G3 (COVID-19 + HIF1A), G4 (recent COVID-19, 2024–2026), G5 (COVID-19 + AI vaccines). Citation counts were obtained from OpenAlex via DOI lookup; for the top-cited papers per group, abstracts were retrieved from PubMed for qualitative review. The full search strategy and executable code are available in the project frameworks cited in Section 2.7 of the main text.

| Group | Query (concise) | N records |
|---|---|---|
| G1 | COVID-19 OR SARS-CoV-2 (MeSH + tiab) | 496,768 |
| G2 | COVID-19 AND (AI OR ML OR deep learning) | 13,545 |
| G3 | COVID-19 AND HIF1A (or HIF-1α) | 139 |
| G4 | COVID-19 papers indexed 2024-01-01 to 2026-06-01 | 104,621 |
| G5 | COVID-19 AND vaccine AND (AI OR ML OR generative) | 1,090 |

*Table S5.1 — PubMed query groups and record counts (2019/01/01–2026/06/01).*

#### S5.2 PRISMA-like flow diagram

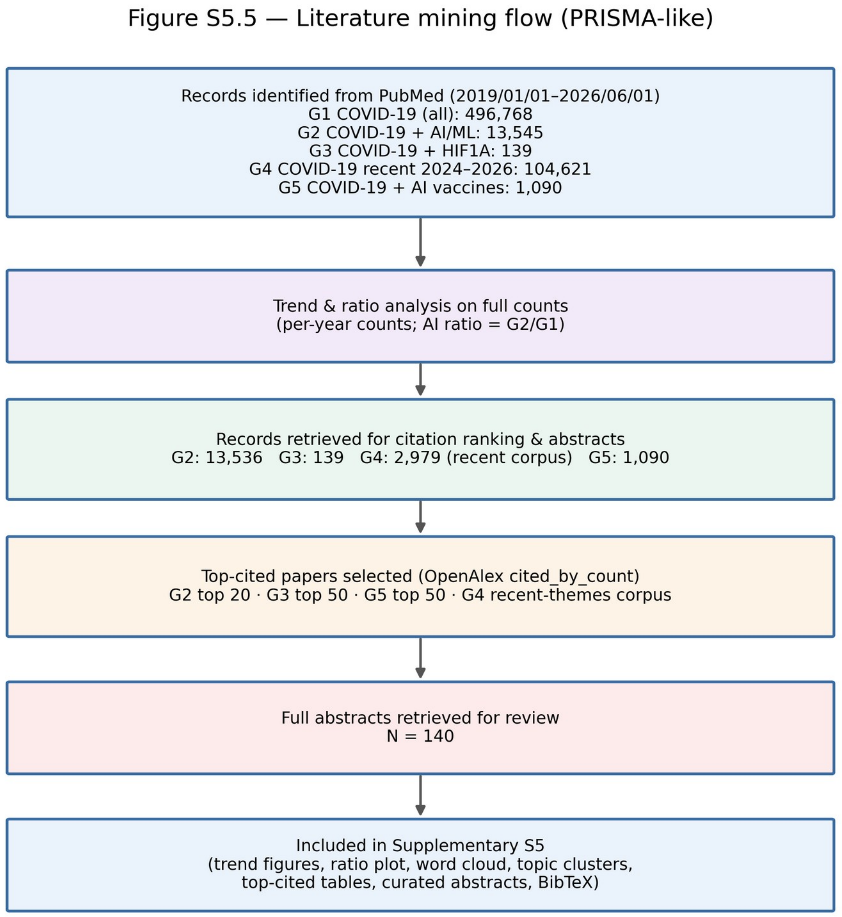


*Figure S5.1 — PRISMA-like flow of the literature mining process: identification (G1–G5), citation ranking, and abstract retrieval.*

#### S5.3 Publication trends and the AI ratio

COVID-19 publication output peaked in 2021 (~138,000) and has gradually declined while remaining substantial: 104,621 papers were indexed across 2024–2026. The AI/ML subset (G2) comprises only 2.70% of all COVID-19 publications across 2019–2026, rising monotonically from 1.45% (2020) to 4.47% (2026 partial). The acceleration aligns with the post-2022 maturation of large language models and other modern AI architectures, indicating that the dominant phase of AI tooling has emerged after the acute pandemic phase and remains under-exploited relative to the corpus size.

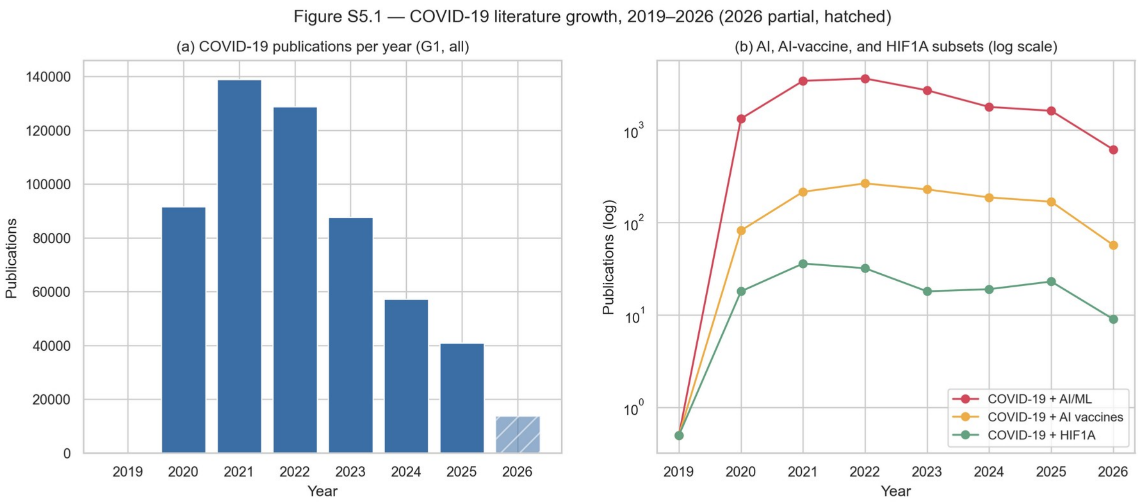


*Figure S5.2 — Annual COVID-19 publications (a) and AI / AI-vaccine / HIF1A subsets on a log scale (b). 2026 is partial (through June 2026).*

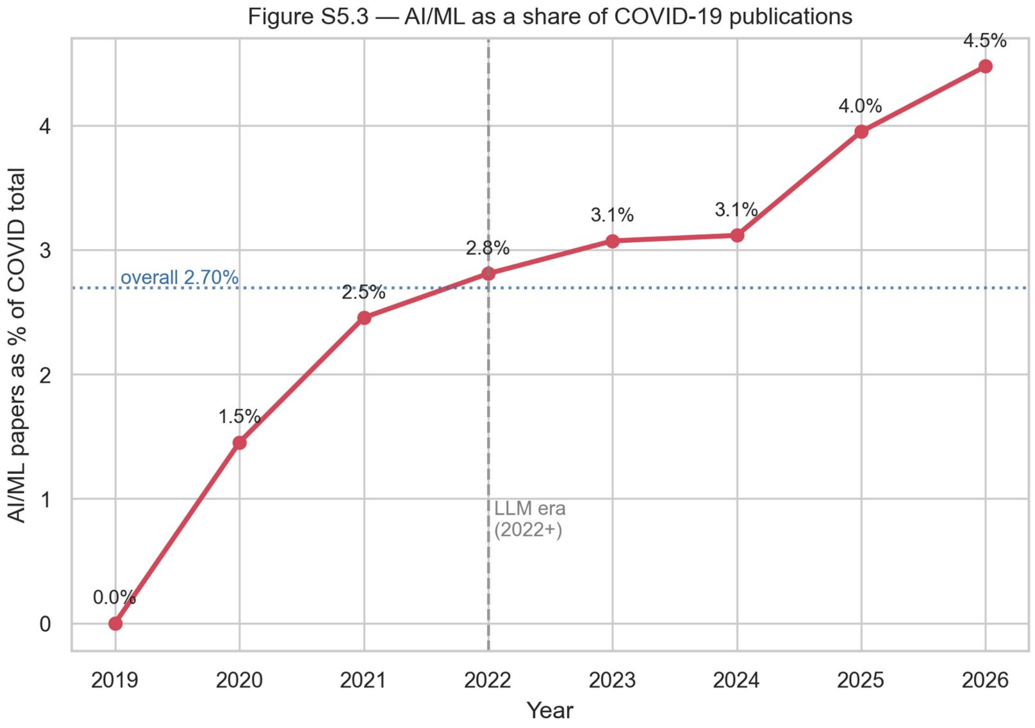


*Figure S5.3 — AI/ML publications as a percentage of COVID-19 publications per year (overall 2.70%); the share accelerates after the 2022 LLM era.*

#### S5.4 Recent themes (2024–2026)

Unsupervised K-means clustering (k=6) of TF-IDF vectors over 2,896 abstracts from the most-cited 2024–2026 papers revealed six themes (cluster shares in parentheses): (i) healthcare systems, workforce, and societal impact (36%); (ii) Long COVID and post-acute clinical outcomes (28%); (iii) molecular virology and host immunity (11%); (iv) mental health (10%); (v) vaccines and mRNA technology (8%); (vi) surveillance and co-circulating respiratory viruses (7%). Cluster (i) mixes in weakly COVID-specific bibliometric noise; clusters (ii)–(vi) carry the substantive recent-research signal. The present HIF1A multi-scale study sits within the molecular virology / host immunity cluster, which remains an active but small thread of contemporary COVID-19 research.

#### S5.5 HIF1A focus and gap analysis

Only 139 HIF1A-related COVID-19 publications were indexed across 2019–2026 — a small niche with no growth trend (annual counts 18–36). The most-cited papers establish the mechanistic HIF-1α ↔ SARS-CoV-2 axis [7-9]. Notably, none of the top-cited HIF1A studies integrate populational, transcriptomic, single-cell, clinical, and host-genetic evidence within a unified predictive framework — the gap the present multi-scale study addresses. Top-cited HIF1A papers are summarized in Table S5.2; full citations and DOIs are provided in the project bibliography.

| # | First author (year) | Journal | Cites |
|---|---|---|---|
| 1 | Codo AC (2020) | Cell Metab | 939 |
| 2 | McElvaney OJ (2020) | Am J Respir Crit Care Med | 673 |
| 3 | Appelberg S (2020) | Emerg Microbes Infect | 300 |
| 4 | Tian M (2021) | Signal Transduct Target Ther | 258 |
| 5 | Serebrovska ZO (2020) | Acta Pharmacol Sin | 219 |
| 6 | Jahani M (2020) | J Inflamm (Lond) | 182 |
| 7 | Guarnieri JW (2023) | Sci Transl Med | 180 |
| 8 | Glushakova OY (2020) | Biomedicines | 167 |
| 9 | Zhu B (2021) | Immunity | 156 |
| 10 | Roncati L (2020) | Ann Hematol | 143 |
| 11 | Bianchi M (2022) | Front Microbiol | 132 |
| 12 | Wing PAC (2021) | Cell Rep | 118 |
| 13 | Saçar Demirci MD (2021) | Gene Rep | 90 |
| 14 | Muhammad Y (2021) | Microvasc Res | 87 |
| 15 | de Cevins C (2021) | Med (NY) | 76 |

*Table S5.2 — Top 15 HIF1A–COVID-19 publications by OpenAlex citation count. Full top-50 list and complete bibliographic data are archived in the project repository.*

#### S5.6 AI-assisted vaccine design outlook

Group G5 (n=1,090) captures heterogeneous applications of AI to COVID-19 vaccine research, ranging from antigen design to vaccine hesitancy text mining. Selected examples of COVID-specific AI vaccine design include reverse-vaccinology approaches [10], deep-learning epitope prediction [11], and mRNA optimization platforms [12, 13]. These COVID-anchored examples complement canonical methodological references (AlphaFold, LinearDesign, ProteinMPNN) cited in the Conclusions of the main text.

| # | First author (year) | Journal | Cites |
|---|---|---|---|
| 1 | Ziegler CGK (2020) | Cell | 2,483 |
| 2 | Vaishya R (2020) | Diabetes Metab Syndr | 1,511 |
| 3 | Antonelli M (2022) | Lancet Infect Dis | 886 |
| 4 | Lalmuanawma S (2020) | Chaos Solitons Fractals | 870 |
| 5 | Chen J (2022) | J Chem Inf Model | 703 |
| 6 | Ton AT (2020) | Mol Inform | 518 |
| 7 | Ong E (2020) | Front Immunol | 453 |
| 8 | Leppek K (2022) | Nat Commun | 373 |
| 9 | Hie B (2021) | Science | 369 |
| 10 | Yang Z (2021) | Sci Rep | 242 |
| 11 | Ong E (2021) | Nucleic Acids Res | 110 |
| 12 | Metkar M (2022) | Acc Chem Res | 108 |

*Table S5.3 — Top AI / COVID-19 vaccine papers by OpenAlex citation count (selected from G5 top-30). The full G5 top-30 list is archived in the project repository.*

*Full data — including all PubMed query results (G1–G5), OpenAlex citation tables, and the curated abstract library for the top-cited papers per group (140 abstracts in total) — are archived in the project repository and available from the corresponding author upon request.*
