## Supplementary material for "Multi-scale analysis of the HIF1A hypoxia–inflammation axis in severe COVID-19": Online Resource 2. Supplementary figures S8-S20

Multi-scale analysis of the HIF1A hypoxia–inflammation axis in severe COVID-19. Journal: Inflammation Research. Authors: Marko N. Živanović, Ivana Raković, Ana Mirić, Miloš Z. Milosavljević, Nevena Milivojević Dimitrijević, Nenad Filipović, Jelena Košarić, Katarina Virijević, Marija Branković, Tamara Mladenović, Biljana Popovska Jovičić. Corresponding author: Marko N. Živanović, Institute for Information Technologies Kragujevac, University of Kragujevac, Serbia;

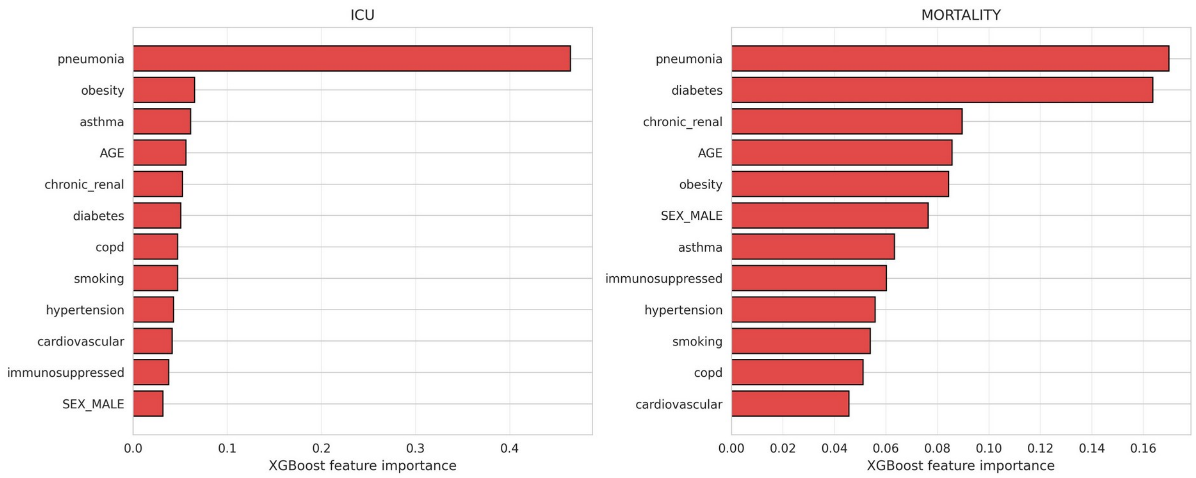


**Fig. S8** Scale A, XGBoost feature importance for ICU admission (left) and death (right) in the Mexican national registry. Radiographic pneumonia dominates both outcomes

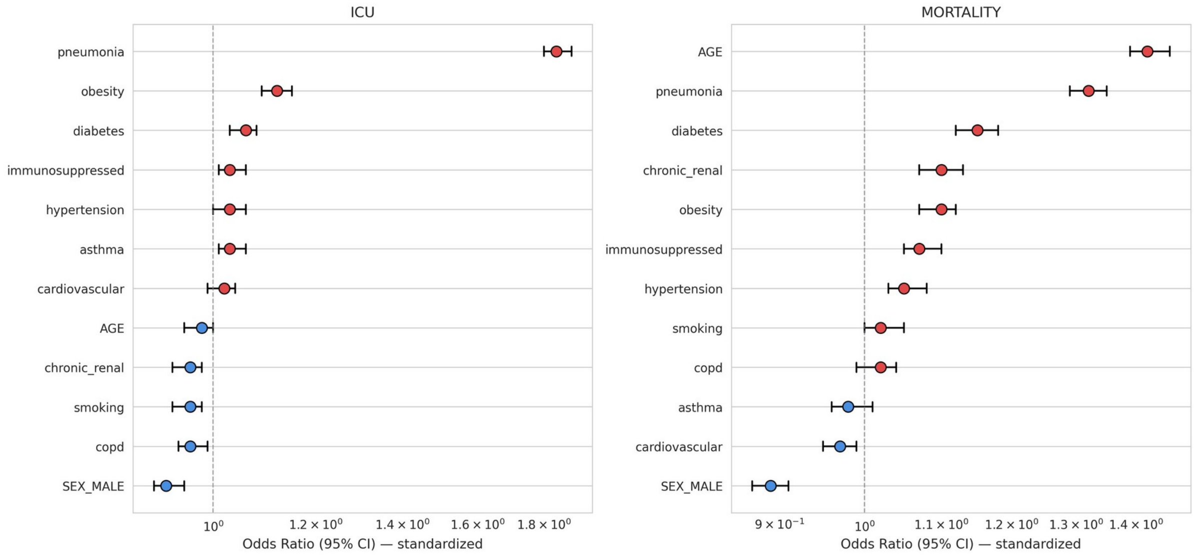


**Fig. S9** Scale A, odds ratios with 95% confidence intervals from logistic regression with standardized predictors for ICU admission (left) and death (right). Male sex shows OR $< 1$ in this registry

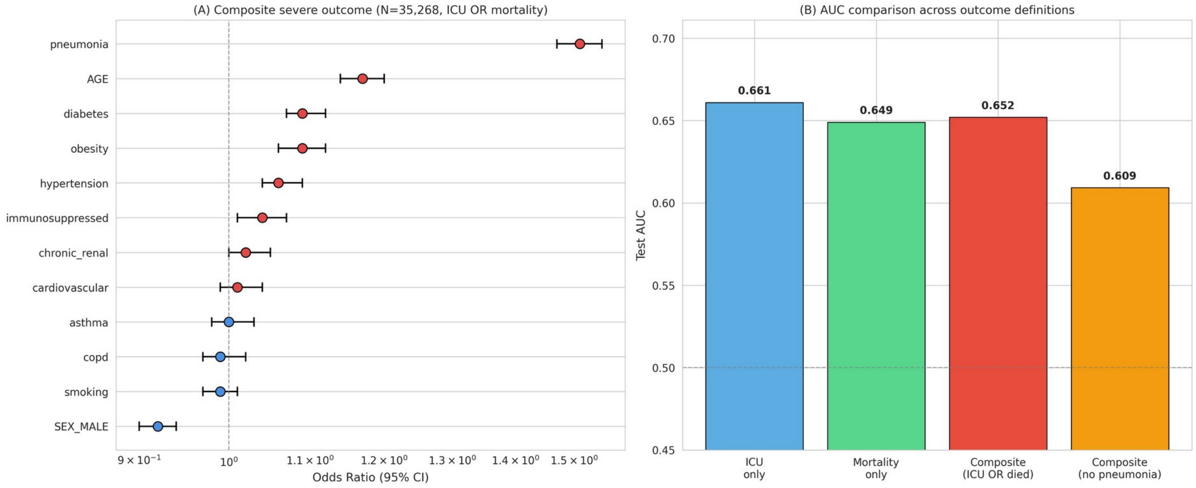


**Fig. S10** Scale A, extensions. Left: odds ratios for the composite severe outcome (ICU admission or death). Right: test-set AUC across outcome definitions, including the composite outcome without the pneumonia variable (AUC 0.609)

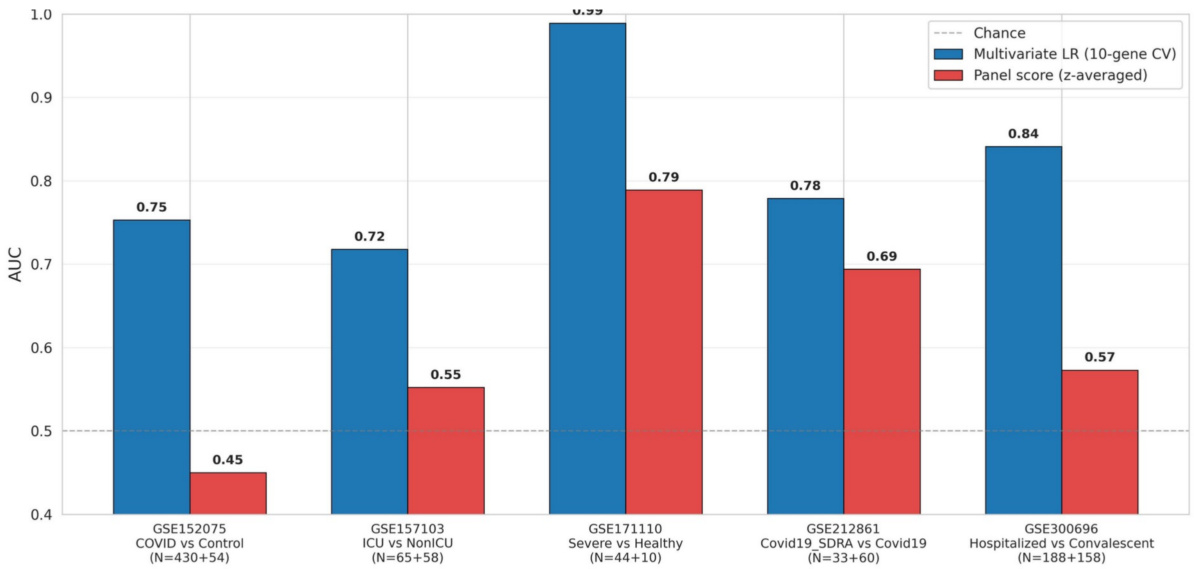


**Fig. S11** Scale B bulk, within-cohort AUC of the 10-gene HIF1A panel: multivariable logistic regression with 5-fold cross-validation (blue) and the unweighted mean of z-standardized genes (red). The gap reflects genes moving in opposite directions within the panel

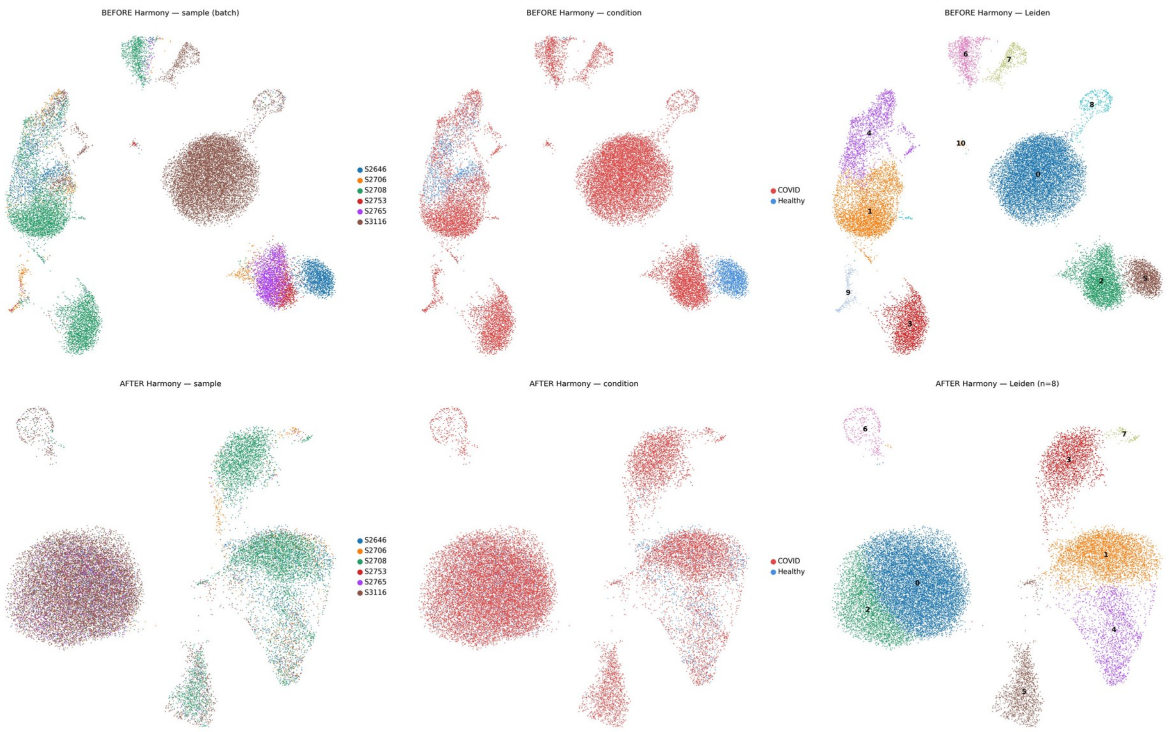


**Fig. S12** Scale B single-cell, Harmony batch integration. Top row, before integration: clusters are dominated by single samples. Bottom row, after integration: samples are interleaved, and maximum single-sample dominance falls to 44–73% for most of the eight Leiden clusters

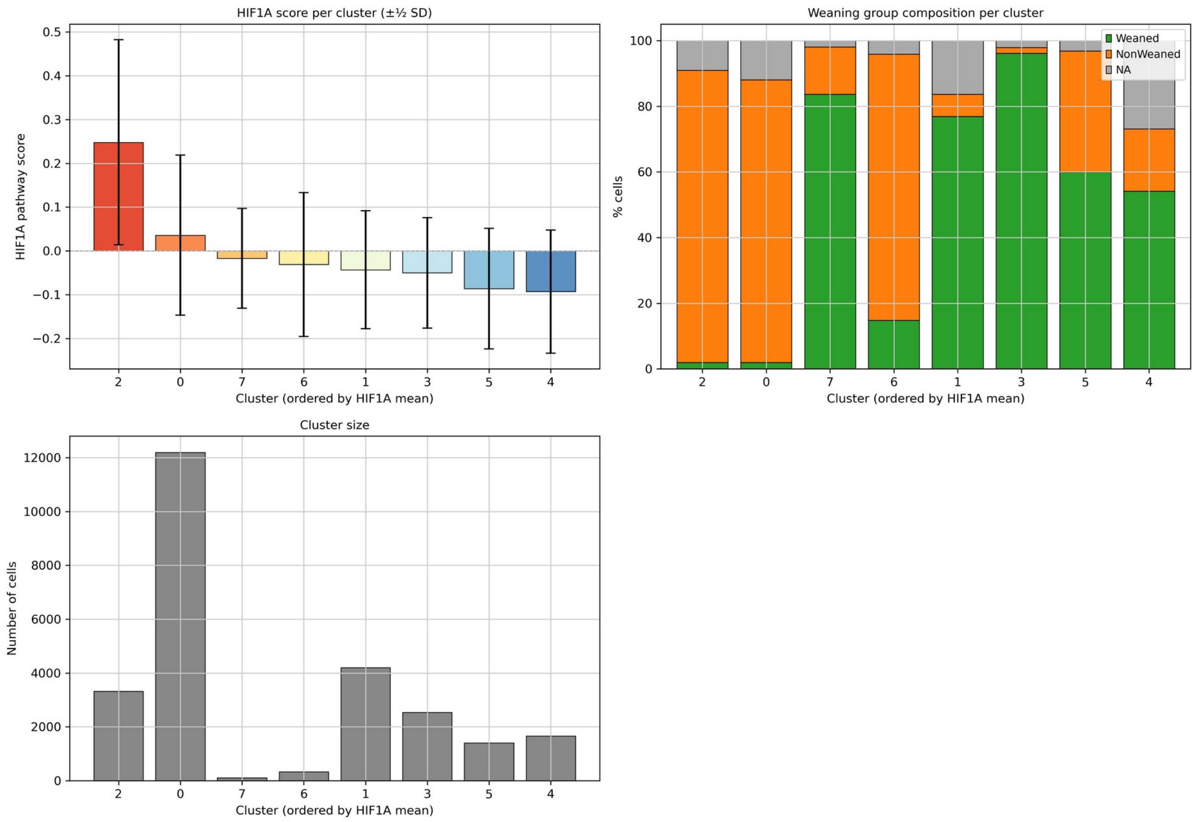


**Fig. S13** Scale B single-cell, per-cluster summary: mean HIF1A pathway score (± ½ SD), weaning-group composition and cluster size. Cluster 2 has the highest score

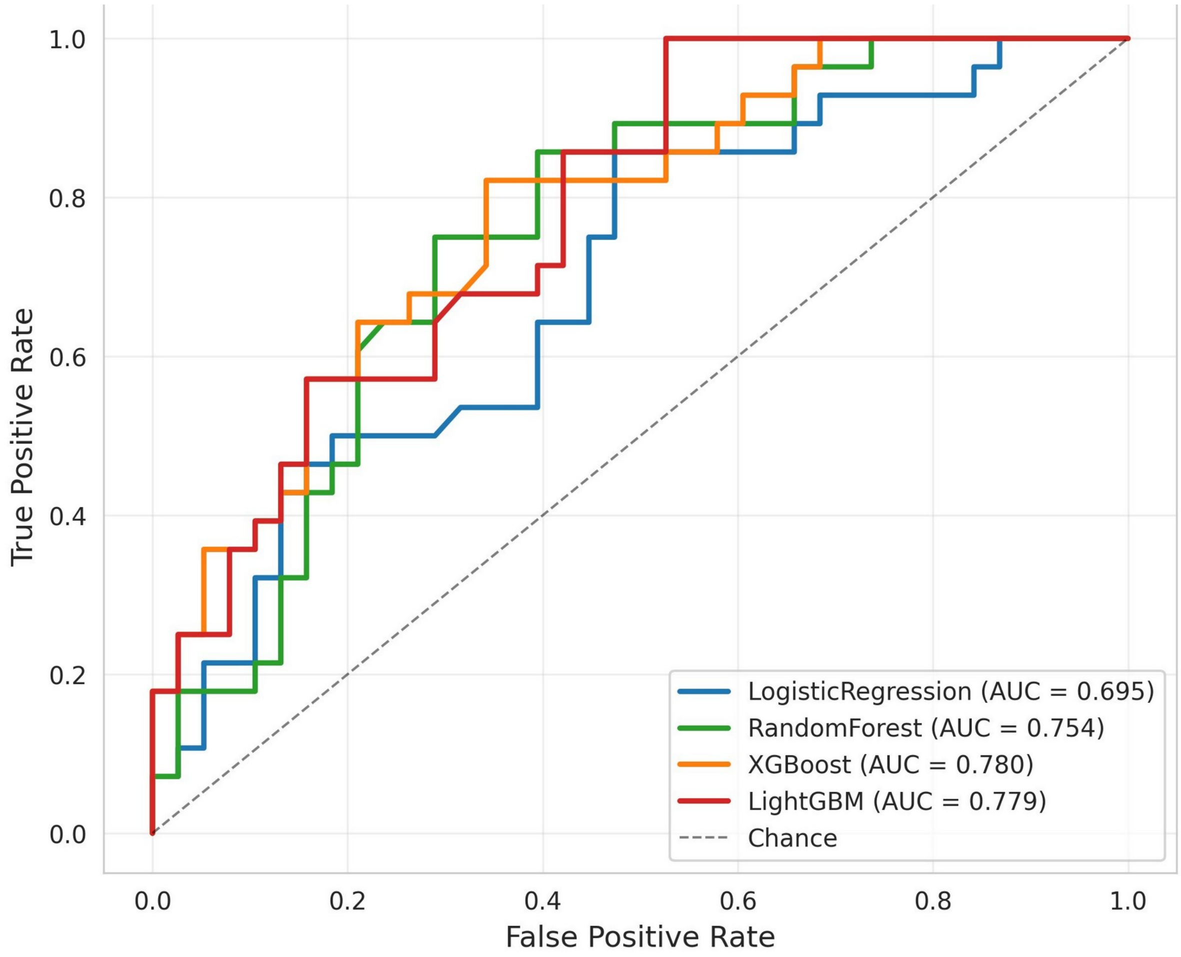


**Fig. S14** Scale C, receiver operating characteristic curves of four models for early ICU transfer on the held-out test set (N = 66)

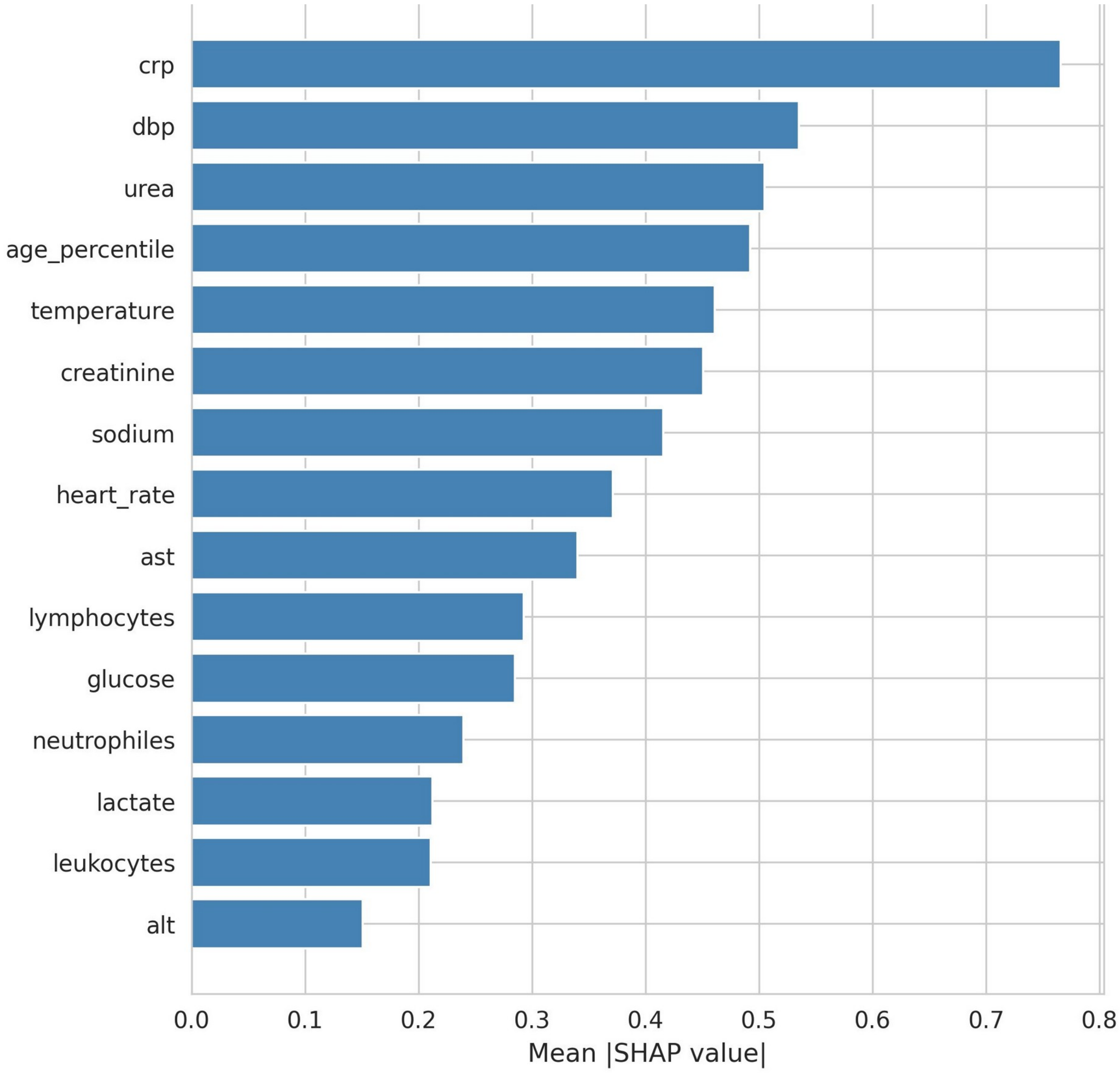


**Fig. S15** Scale C, top 15 features of the calibrated XGBoost model by mean absolute SHAP value

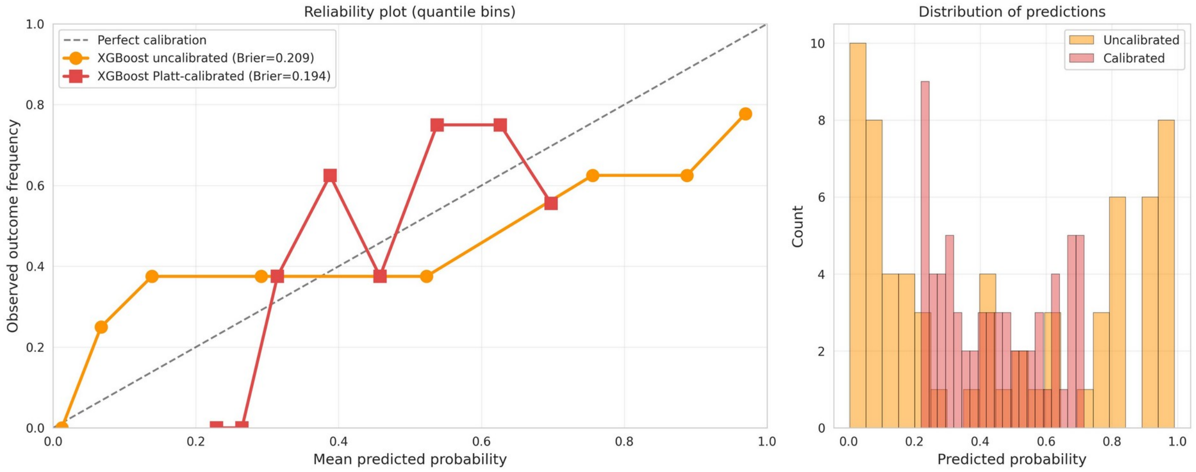


**Fig. S16** Scale C, calibration. Left: reliability plot of uncalibrated (Brier 0.209) and Platt-calibrated XGBoost (Brier 0.194). Right: distribution of predicted probabilities

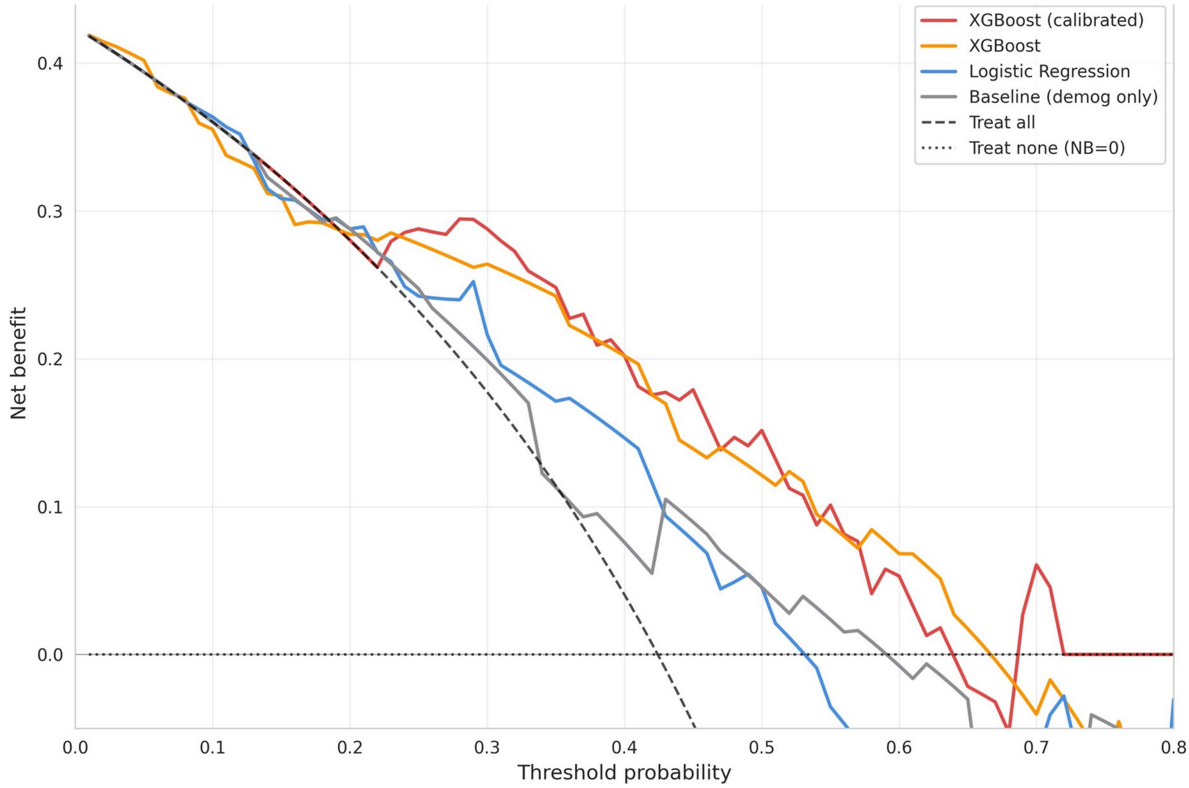


**Fig. S17** Scale C, decision-curve analysis. The calibrated XGBoost model has the highest net benefit across threshold probabilities of 0.14–0.80

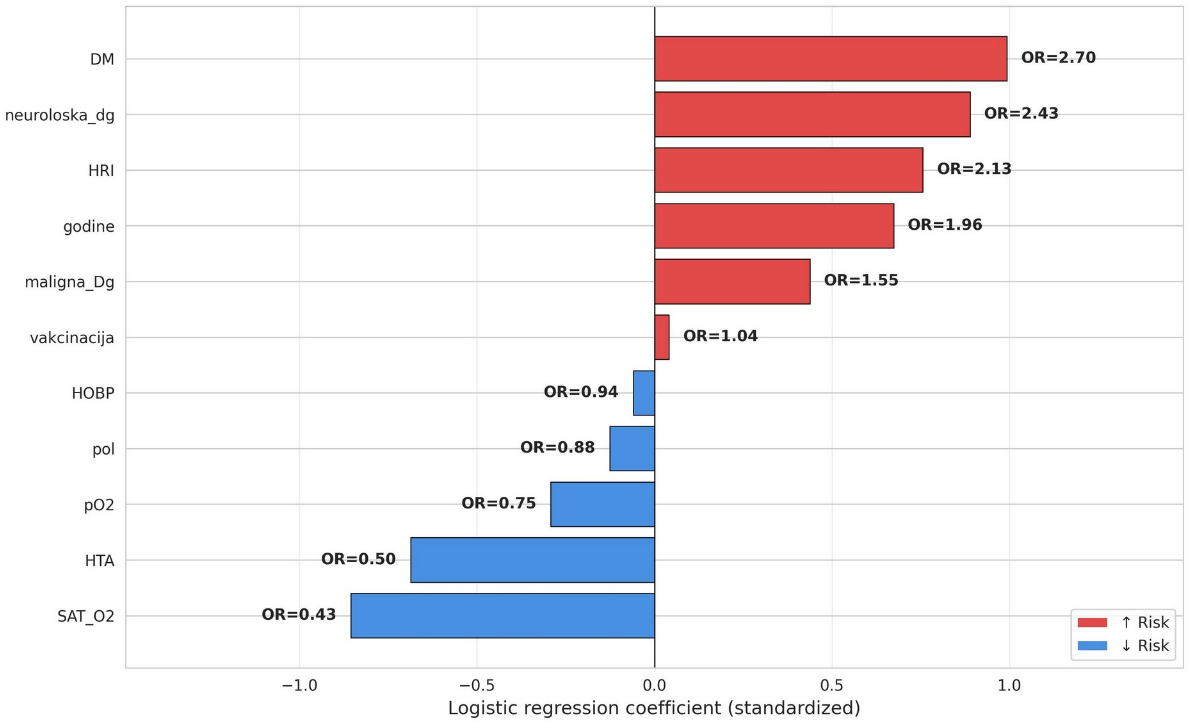


**Fig. S18** Scale D, standardized coefficients of the final L2-penalized, class-balanced logistic regression model for the composite severe outcome (N = 93). Labels: godine, age; pol, male sex; SAT_O2, SpO2; DM, diabetes; HTA, hypertension; HOBP, COPD/asthma; neuroloska_dg, neurological disease; maligna_Dg, malignancy; HRI, chronic renal insufficiency; vakcinacija, COVID-19 vaccination. ORs refer to one standard deviation of each predictor

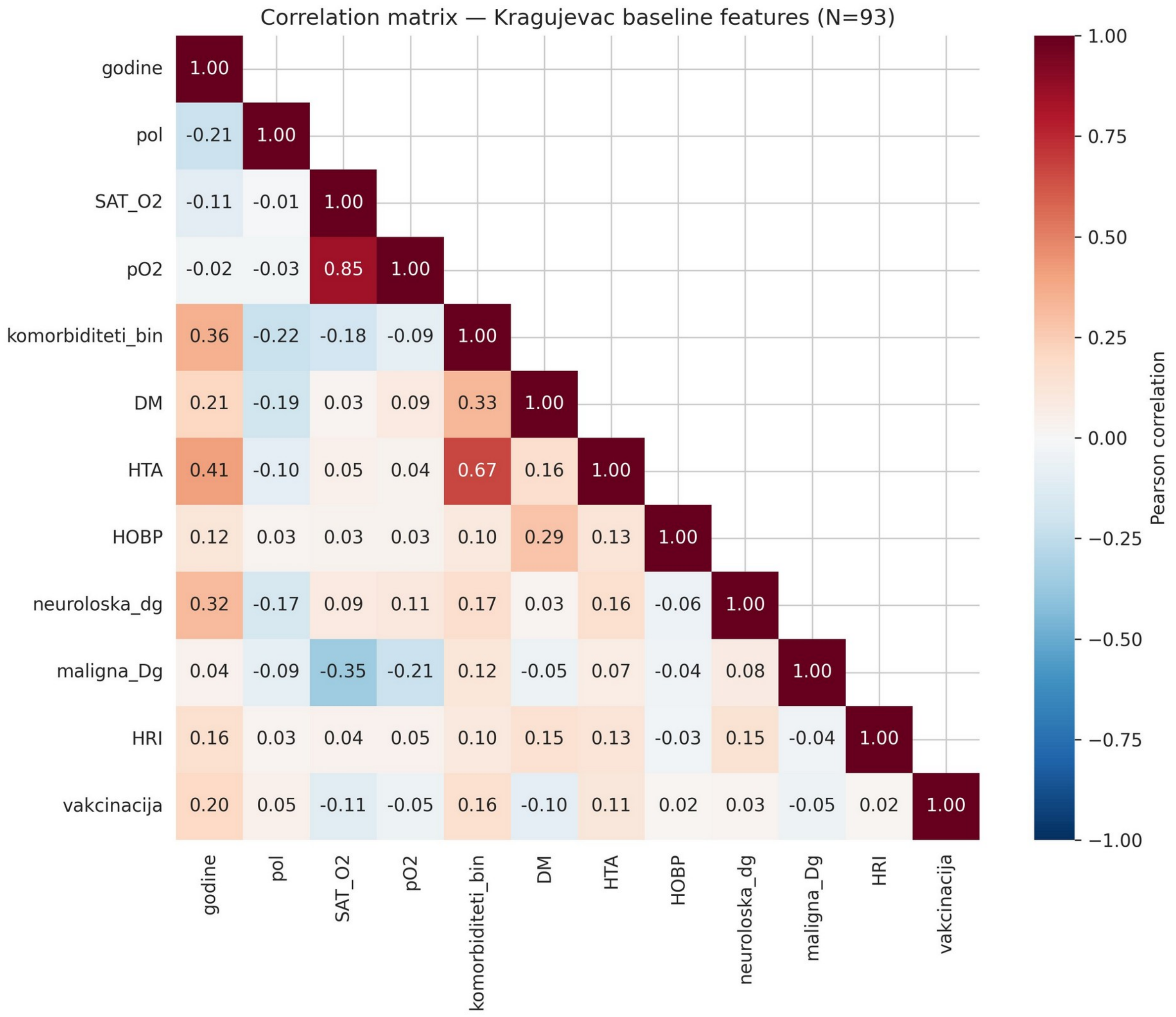


**Fig. S19** Scale D, Pearson correlation matrix of baseline predictors (labels as in Fig. S18; komorbiditeti_bin, aggregated comorbidity indicator, removed from the final model because of collinearity with hypertension). All variance inflation factors of the final predictors were below 5

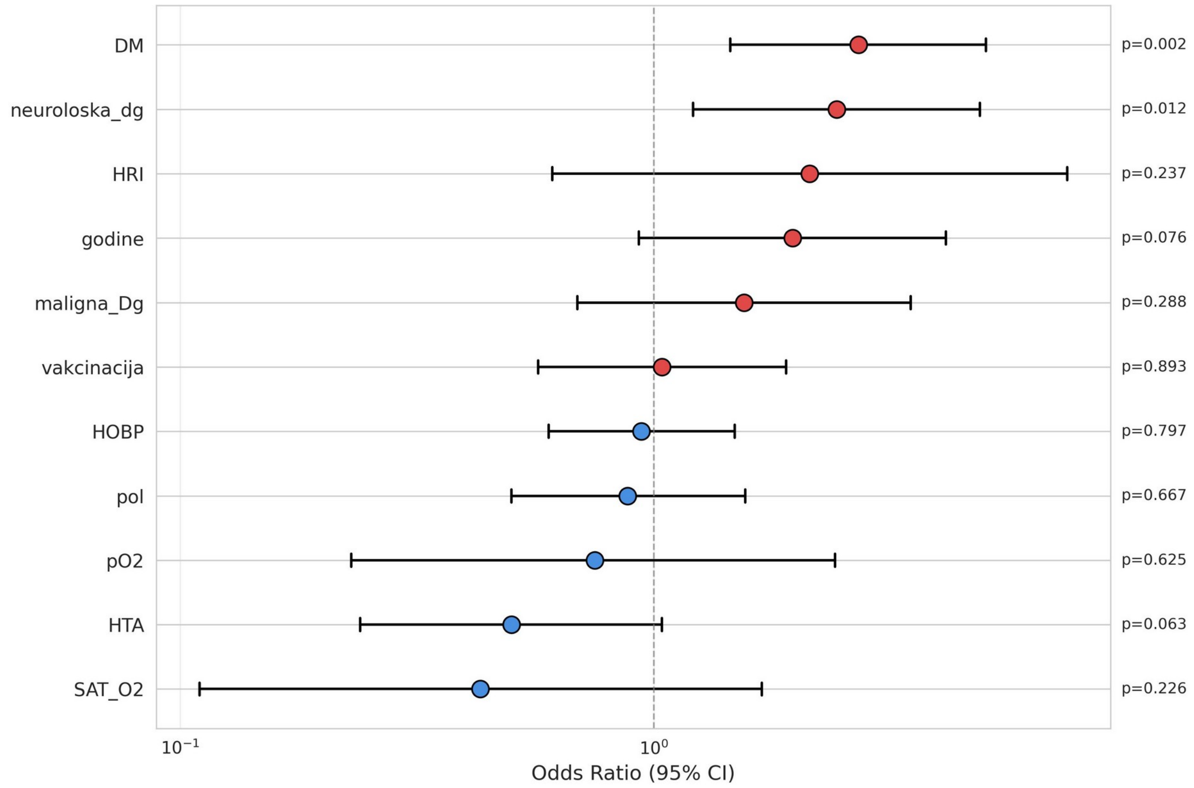


**Fig. S20** Scale D, odds ratios per standard deviation with approximate Wald 95% intervals from the penalized, class-weighted fit (labels as in Fig. S18). Intervals and p values are shown for orientation only and are not used for inference
