## Supplementary material for "Multi-scale analysis of the HIF1A hypoxia–inflammation axis in severe COVID-19": Online Resource 3. TRIPOD checklist

Multi-scale analysis of the HIF1A hypoxia–inflammation axis in severe COVID-19. Journal: Inflammation Research.
Corresponding author: Marko N. Živanović, Institute for Information Technologies Kragujevac, University of Kragujevac, Serbia;. Checklist after Collins et al., BMJ 2015;350:g7594. Sections refer to the main manuscript.

| Item | Section | Checklist item | Where reported / note |
|---|---|---|---|
| 1 | Title | Identify the study as developing and/or validating a multivariable prediction model, the target population and the outcome | Partly. Title names population and pathway; the abstract and Methods identify the prediction models (Scales A, C, D) |
| 2 | Abstract | Summary of objectives, design, setting, participants, sample size, predictors, outcome, analysis, results and conclusions | Abstract (structured) |
| 3a | Background | Medical context and rationale | Introduction, paragraphs 1–3 |
| 3b | Objectives | Objectives, including development and/or validation | Introduction, final paragraph (H1–H4) |
| 4a | Source of data | Study design or source of data | Methods: Scale A, Scale C, Scale D |
| 4b | | Key study dates | Methods: Scale A (2020 registry release); Scale D (enrolment from May 2020). |
| 5a | Participants | Key elements of the setting | Methods: Scales A, C, D |
| 5b | | Eligibility criteria | Methods: Scale A (PCR-confirmed, hospitalized, complete data); Scale C (excluded direct ICU admissions); Scale D (PCR-confirmed, hospitalized) |
| 5c | | Details of treatments received | Online Resource 1, Table S4 (Scale D) |
| 6a | Outcome | Definition of the outcome, including how and when assessed | Methods: Scale A (ICU, death); Scale C (ICU transfer after the first 4 h); Scale D (in-hospital mortality; composite of high-flow oxygen, mechanical ventilation or death) |
| 6b | | Blinding of outcome assessment | Not reported; outcomes were recorded in routine care before the analysis |
| 7a | Predictors | Definition of all predictors, including how and when measured | Methods: Scales A, C, D; Table 2 (admission values) |
| 7b | | Blinding of predictor assessment | Not applicable; predictors were measured at admission, before outcomes occurred |
| 8 | Sample size | How the study size was arrived at | Methods: Statistical analysis (fixed by enrolment; simulated power); Limitations (events per predictor) |
| 9 | Missing data | How missing data were handled | Methods: Scale A (complete-case); Scale D (median imputation) |
| 10a | Statistical analysis | How predictors were handled | Methods: Scale D (standardization; removal of collinear comorbidity indicator, VIF < 5) |
| 10b | | Type of model, model-building procedures and internal validation | Methods: Scale A (LR, RF, XGBoost; cross-validation and held-out test); Scale C (80/20 split, Platt calibration); Scale D (L2-penalized LR, LOOCV) |
| 10d | | Measures to assess model performance | Methods: AUC with bootstrap CI, Brier score, calibration, decision-curve analysis |
| 10e | | Model updating (validation) | Not applicable |
| 11 | Risk groups | Details on how risk groups were created | Scale D: Youden-optimal threshold (Results, Fig. 8) |
| 12 | Development vs validation | Differences from development data in setting, eligibility, outcome and predictors | Results and Limitations: transfer of the Scale C model to Scale D (AUC 0.50) |
| 13a | Participants | Flow of participants, including numbers with and without the outcome | Methods (Scale A filter cascade; Online Resource 1, S1.1); Scale D: 100 enrolled, 93 analysed; Results (events) |
| 13b | | Characteristics of participants | Tables 1 and 2 |
| 14a | Model development | Number of participants and outcome | Methods and Results (all scales) |

| Item | Section | Checklist item | Where reported / note |
|---|---|---|---|
| | | events | |
| 14b | | Unadjusted associations of candidate predictors with outcome | Tables 1 and 2 (Scale D) |
| 15a | Model specification | Full prediction model, including all coefficients and intercept | Partly. Coefficients: Table 3; intercept and fitted model objects: code repository [29] |
| 15b | | Explanation of how to use the model | Not provided; models are exploratory and not proposed for clinical use |
| 16 | Model performance | Performance measures with confidence intervals | Results: Scales A, C, D (AUC with 95% CI, Brier, sensitivity, specificity) |
| 17 | Model updating | Results of model updating | Not applicable |
| 18 | Limitations | Limitations of the study | Discussion: Limitations |
| 19b | Interpretation | Overall interpretation in light of objectives, limitations and other evidence | Discussion |
| 20 | Implications | Potential clinical use and implications for future research | Discussion: Conclusions |
| 21 | Supplementary information | Availability of supplementary resources such as protocol, calculator and data | Declarations: Data and Code availability; Online Resources 1–2 |
| 22 | Funding | Source of funding and role of funders | Declarations: Funding |
